# Microbiome and breast cancer: A systematic review and meta-analysis

**DOI:** 10.1101/2022.12.30.22284053

**Authors:** May Soe Thu, Korn Chotirosniramit, Tanawin Nopsopon, Nattiya Hirankarn, Krit Pongpirul

## Abstract

**Background:** Dysbiosis characterizes breast cancer (BC) through direct or indirect interference in a variety of biological pathways, therefore specific microbial patterns and diversity may be a biomarker for BC diagnosis and prognosis. However, there is still much to determine on the complex interplay of gut microbiome and BC.

**Objective:** To evaluate the microbial alteration in BC patients as compared with control subjects, to explore the gut microbial modification from a range of different BC treatments, and to identify the impact of microbiome patterns on the same treatment-receiving BC patients.

**Methods:** A literature search was conducted using electronic databases such as PubMed, Embase, and the Cochrane Central Register of Controlled Trials (CENTRAL) in *The Cochrane Library* to April 2021. The search was limited to adult BC women and the English language. A prespecified subgroup analysis in BC patients was performed. The results were synthesized quantitatively and qualitatively using random-effects meta-analysis.

**Results:** A total of 33 studies were included in the review, accounting for 20 case-control, 8 cohort, and 5 non-randomized intervention studies. In the meta-analysis, the bacterial DNA load is reduced in the tumor compared with paired normal breast and healthy breast tissue, and interestingly, there is an inverse correlation of the bacterial load in different breast tumor stages. From the intervention studies, it revealed 41 species related to breast tumors with a predominance of *Gemella haemolysans* and *Streptococcus mitis*, and after chemotherapy, the number of species per patient was elevated by a mean of 2.6 (SD = 4.7, p = 0.052). Also, the tumor tissue showed a significant reduction of transcripts of microbial sensors such as TLR2, TLR5, and TLR9, cytoplasmic microbial sensors like NOD1 and NOD2, and the levels of BPI, MPO, and PRTN3. It found that the post-menopausal group has higher leucine-and valine-arylamidase, β-glucuronidase, and esterase-lipase activities in contrast to pre-menopausal and healthy groups.

**Conclusions:** This systematic review elucidates the complex network of the microbiome, BC, and the therapeutic options, expecting to provide a link for stronger research studies and toward personalized medicine to improve their quality of life.

**Funding:** None.

**Registration ID:** PROSPERO 2021 CRD42021288186

## Introduction

Breast cancer (BC) has been of the topmost importance in women for several decades now, with an incidence of 2.3 million in 2020 occupying a prevalence of 11.7% worldwide (1). To date, it has been categorized into four invasive breast cancer subtypes: (1) luminal subtype A, showing a high level of estrogen (ER) and progesterone (PR) receptors, but low expression of human epidermal growth factor receptor (HER2) and cell proliferation index, (2) luminal subtype B revealing ER/PR^+^, HER2^-^ and high proliferation index, (3) HER2^+^ breast cancer subtype, and (4) triple-negative breast cancer (TNBC) subtype (2). Pathological BC staging is critical to figure out how much it spreads in the body and the most common tool is a TNM system in accord to the American Joint Committee on Cancer (AJCC); it stands for tumor (T), node (N) and metastasis (M). According to the AJCC system, effective January 2018, the BC stage for diagnosis and treatment is described by using 7 key pieces of information comprising TNM staging, ER/PR/HER2 status, and cancer grade (3). From the full staging terms of stage I to IV, patients from stage I to IIA are designated as early BC (3, 4).

Typically, multiple risk factors are increasing the possibility to occur breast cancer such as aging, sex, obesity, family history, genetic mutation, estrogen level, and sedentary lifestyles (5-7). In contrast, the human microbiota raised so much attention as a major risk modulator due to their distinct role in the regulation of steroid hormone metabolism, through the stimulation of different enzymes, such as hydroxysteroid dehydrogenase (8, 9). Normally, the microbiota is in homeostasis in the human host, affecting local and distant organs (10). With unfavorable conditions, the community is imbalanced, then leading to promote many health issues (11-13).

It is also evident that relative species abundance is not different between normal women and premenopausal patients but found significant differences in postmenopausal patients in whom estrogen is non-ovarian (14). In addition, the microbiota secretes bioactive bacterial metabolites that can modulate the disease progression, and these are reactivated estrogens, amino acid metabolites, short-chain fatty acids (SCFAs), or secondary bile acids (BAs) (15-17). Therefore, the impact of the intestinal microbiome is multi-factorial, and it is critical in regulating the host immune system in the pathophysiology of BC development and therapy response and/or resistance (18).

So far, the gut microbiome explains different responses to a variety of cancer therapies (19-21). In animal models, it revealed that the gut microbiota is modulated by chemotherapy leading to side effects of early BC treatment like weight gain or neurological abnormalities (22). The experimental studies also demonstrated the correlation of the intestinal microbiota in BC subtypes with the clinical outcomes and therapeutic response (23, 24). Particularly, a study discovers that gut bacteria are superabundant in BC patients in contrast to healthy individuals and hurt BC prognosis (22). Therefore, the comprehensive insights into the oncobiome of BC subtypes are significant for therapeutic purposes and disease prognosis, however, additional research is required to discover their complexity for the BC prognosis.

At the gene level, multiple genes that metabolize estrogens and different enzymes like β-glucosidase and glucuronidase have been explored (25, 26). Since the intestinal microbes encode the genes, their prominent role is in the regulation of steroid hormone metabolism (27, 28) and subsequently, these endogenous estrogens are critical in the BC progression (29-31). The fecal β-glucuronidase activity is negatively related to estrogen levels in the gastrointestinal system, and more importantly, the richness of the fecal microbiome is directly and markedly correlated with systemic estrogen levels (8).

In the past decades, numerous studies revealed the impact of the microbiome on different organ-specific cancers and the action of bacterial metabolites in the human host in several signaling pathways, e.g., E-cadherin/β-catenin pathway, breaking DNA double-strands, promoting apoptosis, and altering cell differentiation (32-34). Particularly, there still are several questions between the human microbiome and BC development ‘What pattern of the microbiome profile the breast cancer patients have in contrast to non-breast cancer subjects’; ‘How the microbiome is being modified by different treatments’; and ‘What are the microbiome profile on the same treatment’. To address these, a systematic literature review and meta-analysis on breast cancer and microbiome are conducted, and the specific objectives are to evaluate the microbiota alteration in breast cancer patients compared to non-breast cancer subjects, to explore the microbiota modification from a range of different treatment strategies, and to identify the impact of microbial pattern on the same treatment-receiving breast cancer patients.

## Materials and Methods

### Protocol and registration

The systematic literature review was registered on PROSPERO ID 2021 CRD42021288186.

### Literature search

The PRISMA statement guidelines were followed to conduct the systematic literature review and meta-analysis (35). The inclusion and exclusion criteria for the study were based on PICO/PECOs framework (36, 37). Two of the authors (MT, KC) independently examined each study for inclusion in the systematic review by using PubMed, Embase, and the Cochrane databases, and discrepancies were resolved through group discussion at every step. The search was limited to adult BC women and the English language. Studies that included only non-human subjects or were not peer-reviewed were excluded. Both epidemiologic and intervention studies were considered from these databases and mainly focused on the interlink between BC patients and intestinal microbiome that was being extracted to April 2022.

### Study selection

Article screening was done by two independent reviewers (MT, KC) for eligible studies using pre-specified inclusion and exclusion criteria, followed by the full-text reviewing process. All the relevant full-text papers were taken for further data extraction. The inclusion criteria for the meta-analysis were set up as follows: 1) epidemiologic studies on how microbiome profile in BC patients differed from the pattern in non-BC control, and 2) intervention studies of how the treatment in BC patients affected the microbiome and vice versa. The exclusion was performed on 1) the studies such as animal studies, *in-vitro*, review articles, non-peer-reviewed articles, protocols, letters, editorial, commentary, recommendations, and guidelines; and 2) the studies on BC survivors. Disagreements between the review authors were resolved by consensus at every phase of the systematic review screening.

### Data extraction

The independent two authors (MT, KC) performed the data extraction for the following variables: 1) Authors, year of publication, study period, study type, and the country which implemented the study; 2) Demographics characteristics such as menopause, menarche, and hormonal status; 3) Related characteristics including the cytokine levels and enzymatic activities; and 4) Parameters for diversity profile. All relevant text, tables, and figures were examined during the data extraction, and discrepancies between the two authors were resolved by discussion or consensus.

### Risk of bias

The independent two authors (MT and KC) assessed the risk of bias (ROB) in the extracted intervention studies. However, the studies are non-randomized trials, and therefore ROBINS-I (Risk Of Bias In Non-randomized Studies - of Interventions) tool was applied to assess ROB (39). For included cohort and case-control studies, the two independent authors (MT and KC) performed the ROB assessment using the Newcastle - Ottawa Quality Assessment Scale (NOS) that was developed from an ongoing collaboration between the Universities of Newcastle, Australia, and Ottawa, Canada for quality assessment in a meta-analysis (40). The tool was used to assess the following domains: bias arising from the selection process; bias arising from the comparability process; and bias arising from the outcome/exposure process.

For any disagreement, it was resolved by consensus. If there were not enough information to consider, the corresponding authors were emailed, and their response was waited for two weeks. In the case of no response, it proceeded with the available data, and any disagreement was resolved through discussion.

### Subgroup analysis

The analysis was conducted in the following subgroups: Menopausal status including pre-menopause, peri-menopause, and post-menopause; Menarche type; Hormonal status accounting for ER, PR, HER-2, and CerB-2; Sample type; and BC stage.

### Statistical analysis

For intervention studies, mean differences (MD) along with a 95% confidence interval (95% CI) between groups were indicated for microbiome diversity outcomes. Participant characteristics, study period, study type, and study location were assessed for clinical and methodological heterogeneity. The *I*^*2*^ and *X*^*2*^ statistics were used for the assessment of statistical heterogeneity. The heterogeneity level was as defined in Chapter 9 of the Cochrane Handbook for Systematic Reviews of Interventions and the *X*^*2*^ test was assessed with a p-value of less than 0.05 meaning as statistically significant. For clinical, methodological, and statistical heterogeneity, the random-effects meta-analysis by DerSimonian and Laird method was utilized by RevMan 5 software.

## Results

### Study selection

The literature search found 2,761 articles from the databases of which 758 duplicates were removed before the screening. From the initial 2,003 studies, the title and abstract screening were performed, and 1,884 articles were excluded according to the inclusion and exclusion criteria. Next, we retrieved 119 articles for the full-text screening and checked their eligibility for the meta-analysis. Amongst, 86 studies were excluded due to the following conditions: 50 studies are non-peer reviewed articles, 11 targeted the wrong population, 7 raised wrong outcomes, 4 were protocol papers, 3 were editorial, 3 were wrong intervention, 3 were wrong study design, 1 were wrong comparator, 2 were review articles, 1 was a duplicate, and 1 were not reported in English. Lastly, 33 studies with an enrollment of 3,544 participants covering the study period from 2004 to 2019, were included in the systematic literature review and meta-analysis, accounting for 20 case-control, 8 cohort, and 5 non-randomized intervention studies (Figure 1).

**Figure 1.**
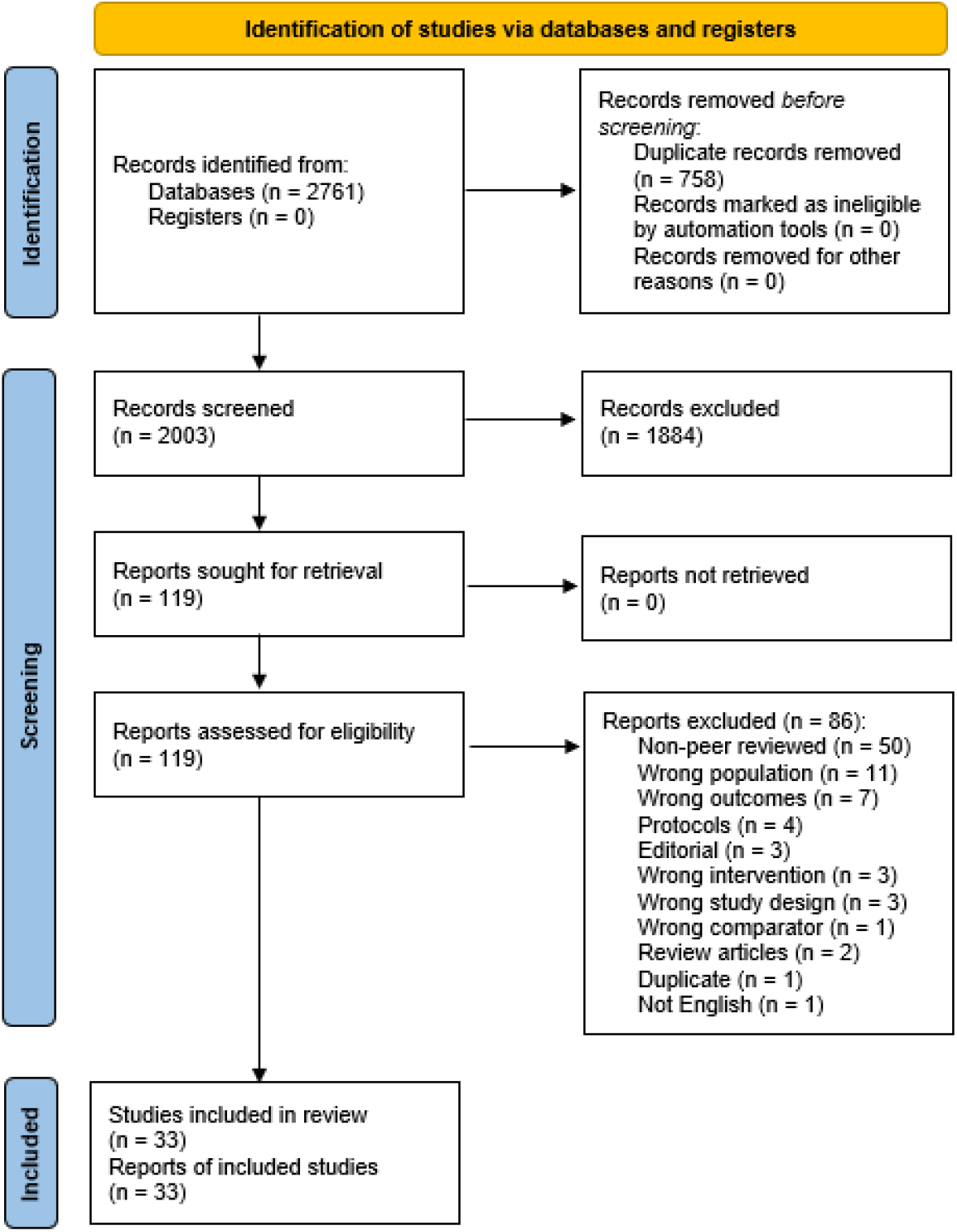
Flow diagram of the study selection in the systematic literature review

### Study characteristics

The extracted studies were published from 1990 to 2020 in 11 countries, mostly contributed from USA and China. It included different study types such as cohort, case-control, and intervention studies, and the participated age range was 18-90 years (Table 1).

**Table 1.** Baseline characteristics of the included studies

| First Author | Published year | Study period | Study type | Country | Participants, n | Age Range (yr) | Ref. |
| --- | --- | --- | --- | --- | --- | --- | --- |
| Minelli | 1990 | - | Case-control | Italy | 48 | 25-52 | (42) |
| Benini | 1992 | - | Case-control | Italy | 73 | 25-52 | (43) |
| Xuan | 2014 | - | Case-control | USA | 20 | - | (44) |
| Urbaniak | 2014 | 2012 | Case-control | Canada & Ireland | 81 | 18-90 | (45) |
| Goedert | 2015 | - | Case-control | USA | 96 | 50-74 | (46) |
| Banerjee | 2015 | - | Case-control | USA | 137 | - | (47) |
| Urbaniak | 2016 | - | Case-control | Canada | 71 | 19-90 | (48) |
| Hieken | 2016 | - | Case-control | USA | 33 | 33-84 | (49) |
| Wang | 2017 | 2014-2016 | Case-control | USA | 78 | - | (50) |
| Thompson | 2017 | - | Case-control | USA | 740 | - | (51) |
| Goedert | 2018 | - | Case-control | USA | 96 | 50-74 | (52) |
| Huang | 2018 | 2006-2015 | Case-control | Taiwan | 5 | - | (53) |
| Banerjee | 2018 | - | Case-control | USA | 168 |  | (54) |
| Zhu | 2018 | - | Case-control | China | 133 | - | (14) |
| Smith | 2019 | - | Case-control | USA | 83 | 18-72 | (55) |
| Ma | 2020 | 2017-2018 | Case-control | China | 50 | - | (56) |
| Klann | 2020 | - | Case-control | Switzerland | 46 | - | (57) |
| UzanYulzari | 2020 | - | Case-control | Israel | 33 | 18-75 | (58) |
| He | 2021 | 2019 | Case-control | China | 82 | 18-49 | (59) |
| Byrd | 2021 | - | Case-control | Ghana | 895 | 18-74 | (60) |
| Luu | 2017 | - | Cohort | France | 31 | 39.6–79.3 | (61) |
| Meng | 2018 | - | Cohort | China | 94 | 29-77 | (62) |
| Costantini | 2018 | - | Cohort | Italy | 16 | 46-82 | (63) |
| Shi | 2019 | 2017 | Cohort | China | 80 | <45, 45-59, ≥60 | (64) |
| Yoon | 2019 | 2016-2017 | Cohort | Korea | 121 | 32-78 | (65) |
| Thyagarajan | 2020 | - | Cohort | USA | 23 | 27-78 | (66) |
| DiModica | 2021 | 2017-2019 | Cohort | Italy | 24 | - | (67) |
| Yao | 2020 | 2019 | Cohort | China | 36 | - | (68) |
| Napeñas | 2010 | 2004-2006 | Non-randomized trials | USA | 9 | 33-69 | (69) |
| Frugé | 2020 | 2014-2017 | Non-randomized trials | USA | 32 | - | (70) |
| Chiba | 2020 | 2004-2014 | Non-randomized trials | USA | 42 | - | (71) |
| Wu | 2020 | - | Non-randomized trials | USA | 37 |  | (72) |
| Guan | 2020 | - | Non-randomized trials | China | 31 | 36-66 | (73) |

### Subject characteristics

Overall, the combined mean of the participants’ age is 55.07 with a standard deviation of 5.46. Regarding the menopausal status of the participants, there are 3 main groups: pre-menopausal, peri-menopausal, and post-menopausal subjects. Amongst 66.4% of the breast cancer cases are post-menopause patients, and 46% of the cancer patients are ≥13 years of age at menarche. In the study, 54% of the breast cancer patients are from the USA, followed by Ghana with 15% and China with 14% (Table 2).

**Table 2.**
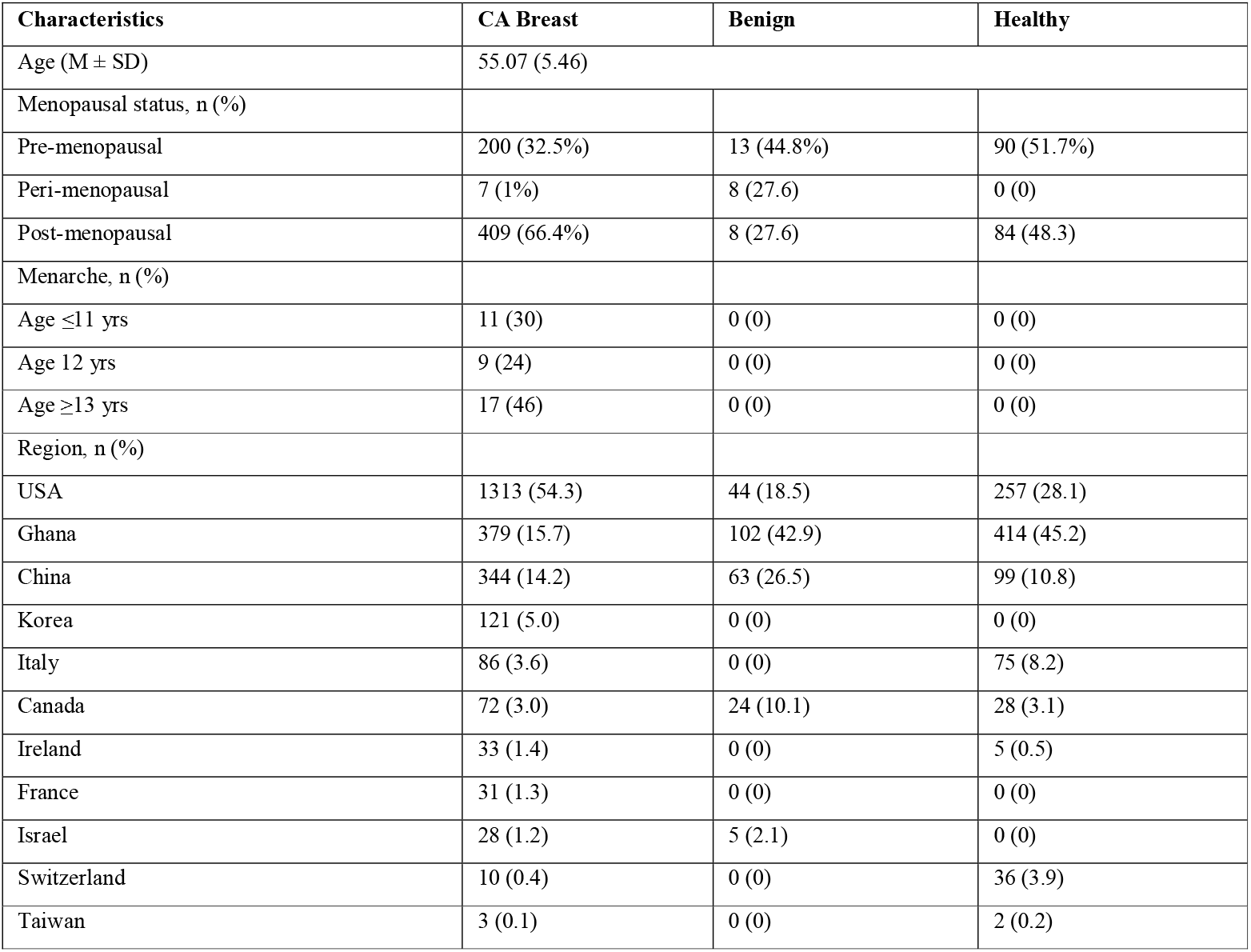
Demographic characteristics of participants

### Risk of bias

The ROB of the included studies was summarized by the study design group. The ROB on case-control studies was assessed mainly on the categories of selection, comparability, and exposure (Figure 2). In eleven studies, they followed to collect the cases with independent validation while the others accumulated the case population based on archived medical records or databases. Regarding the representativeness of the cases, 4 studies had the potential for selection biases at which one study used the subjects from their Army, and another study took the samples from the genomics data repository with no clear statement for selection. Mostly the control subjects were obtained as hospital controls, for example, women who underwent breast reductions or cosmetic procedures, and control subjects from 7 studies were recruited from the community. Amongst, 2 studies used benign cases as controls and 1 study put the other malignancy cases as controls. All the cases and control subjects had comparability based on the study design and the additional comparability measures were the same Mediterranean diet, age, and hormonal status by menopause. The exposure was mostly known by surgical records, followed by written medical records. All the subjects’ selection was followed by the same methods of ascertainment and the same rate for both groups.

**Figure 2.**
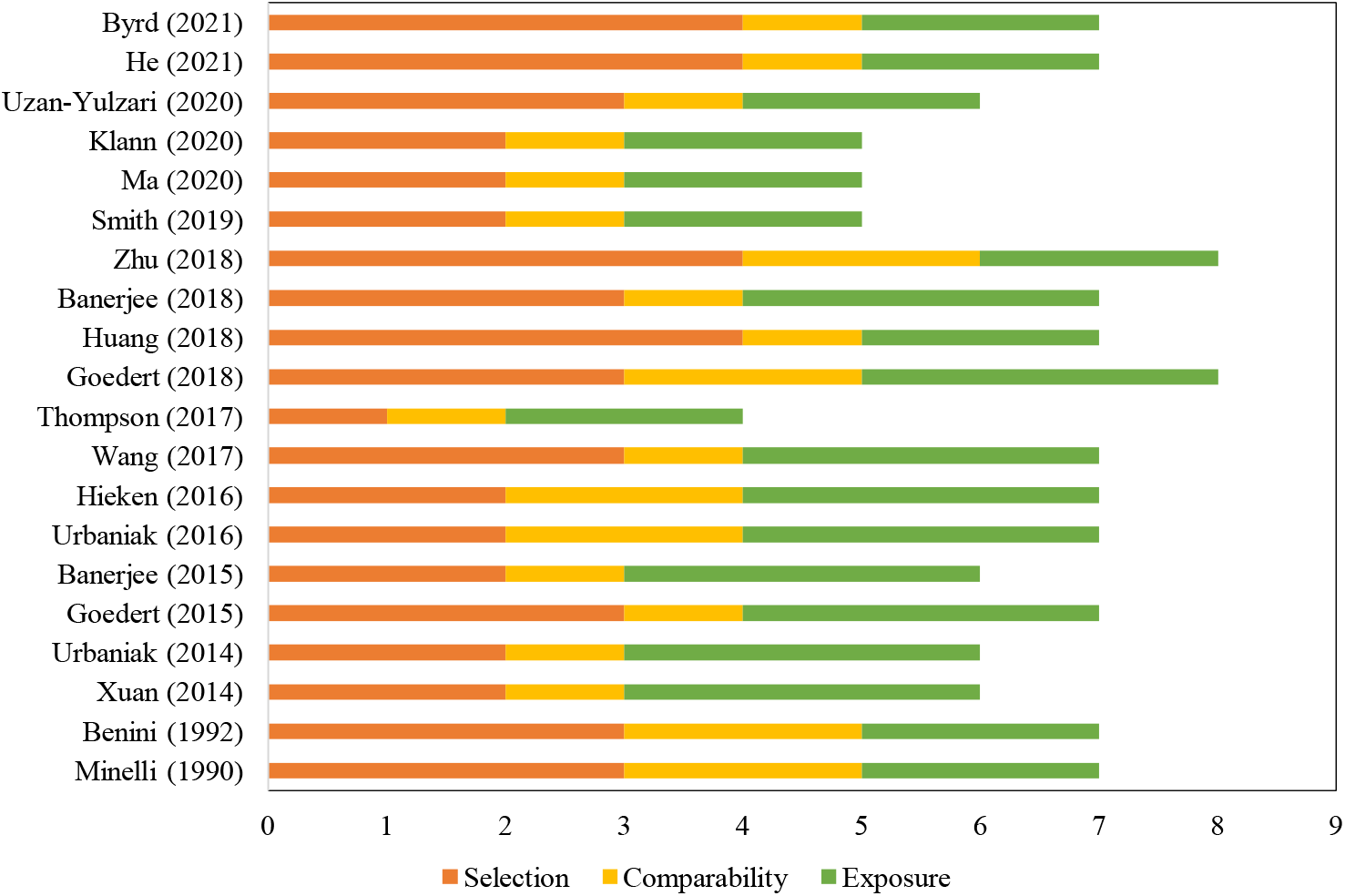
Risk of bias of the included case-control studies via NOS scale

The ROB on 8 cohort studies was assessed by the selection, comparability, and outcome (Figure 3). The selection of exposed cohorts was truly or somewhat representative of the community in which 1 study used people from a volunteer group that was not representative of the community. And all studies had a comparability of cohorts based on the study design. The ascertainment of outcomes was from medical records with probably enough follow-up time, however many studies did not describe any statement for the adequacy of the follow-up at the cohorts. In a study, some subjects were excluded from the final analysis because of admixed ancestry.

**Figure 3.**
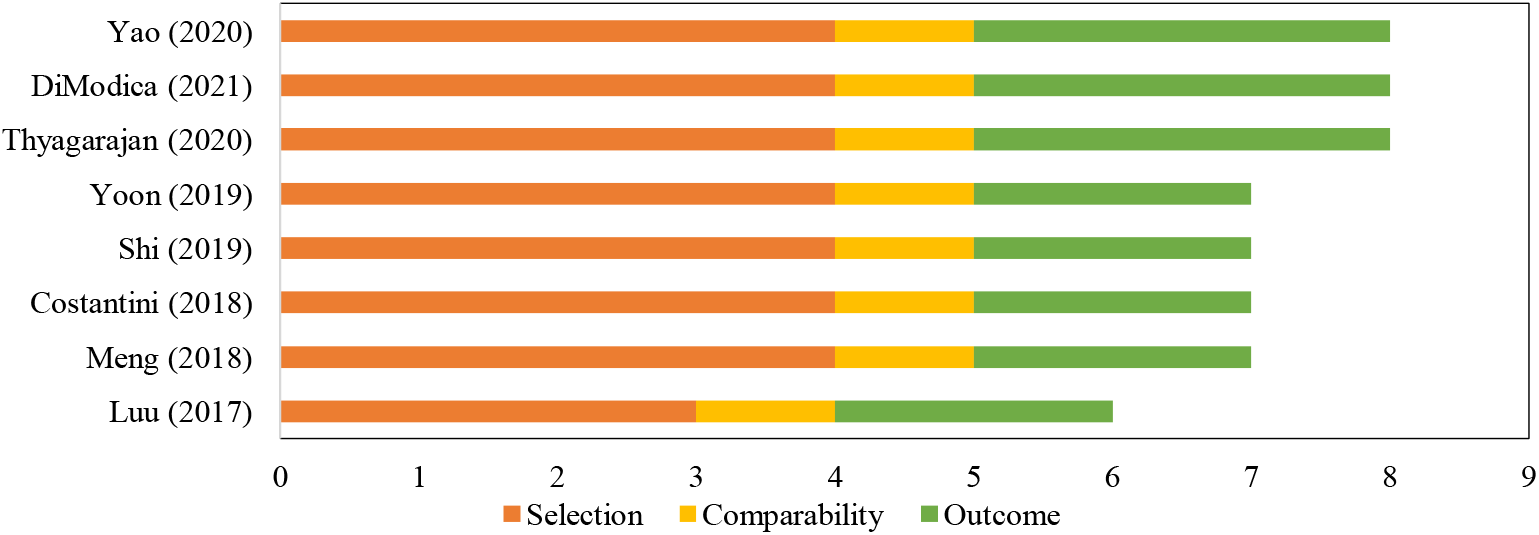
Risk of bias of the included cohort studies via NOS scale

At the ROB assessment of non-randomized intervention trials (Figure 4), most study domains were recognized as low risk of bias. Four articles did not describe the process of participant selection. In the study by Fruge et al. (2020), there was a bias of confounding. The study by Wu (2020) reflected a deviation bias from intended interventions.

**Figure 4.**
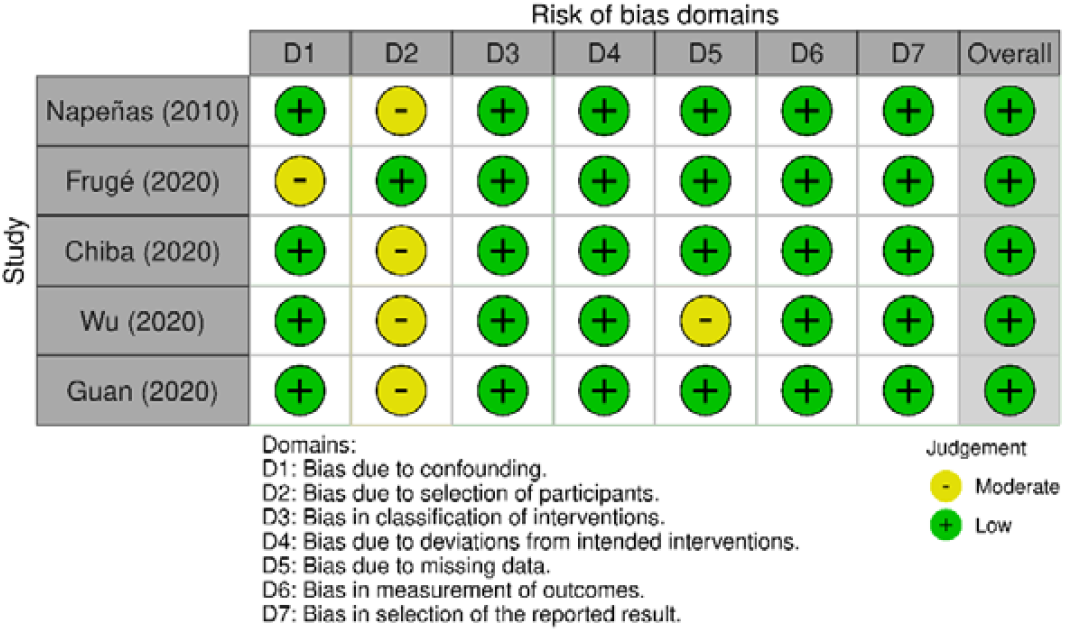
Risk of bias assessment for non-randomized intervention studies

### Qualitative analysis

Generally, there were numerous qualitative data while analyzing the microbial communities from different studies due to different sampling areas, several sequencing techniques, and diverse geographical and biological conditions. Therefore, all the extracted studies were qualitatively analyzed by groups of case-control, cohort, and non-randomized intervention studies (Table 4-6), accounting for the study period, sample information, mean age of the population, microbial detection methods, bacterial profile, and summary of the studies.

### Risk of breast cancer by the microbiome

An overall significant reduction of observed species for the BC group was found in 2 studies (MD = -20.16; 95% CI = -34.66 to -5.66; p = 0.006), however, the heterogeneity value is high (*I*^*2*^ = 87%; p = 0.006) (Figure 5).

**Figure 5.**
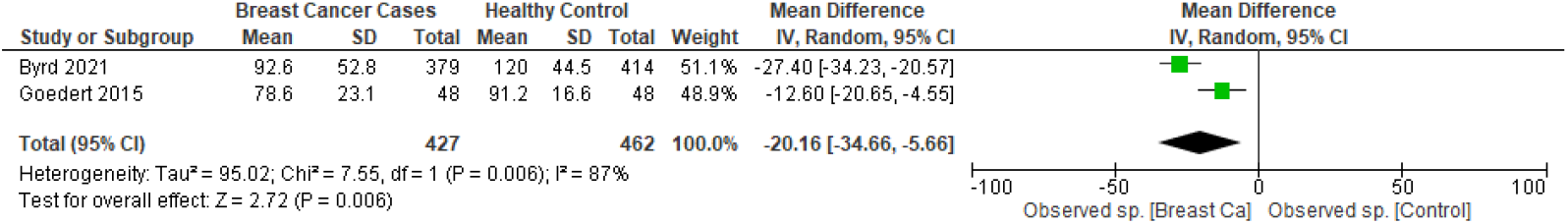
Meta-analysis forest plot representing the risk of breast cancer by observed species of microbial profiling.

The overall estimates of the Shannon index for the BC group reported a significant reduction compared with healthy subjects even though the study by Goedert (2015) was not significant (MD = -0.35; 95% CI = -0.48 to -0.22; p < 0.00001) (Figure 6). The heterogeneity between studies was moderate (*I*^*2*^ = 41%; p = 0.19).

**Figure 6.**
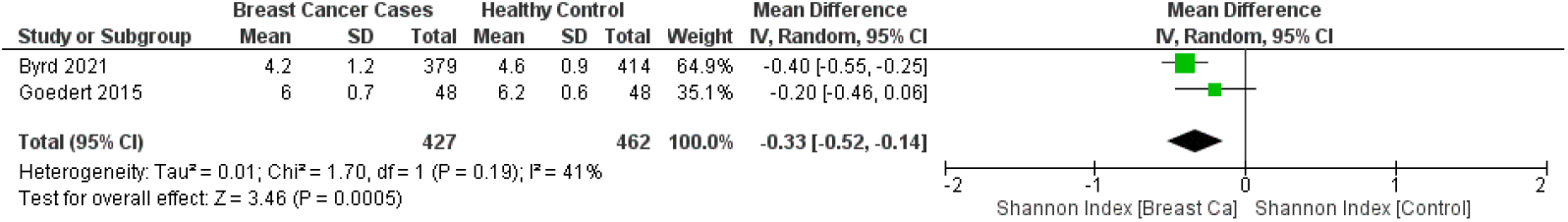
Meta-analysis forest plot representing the risk of breast cancer by Shannon index of microbial profiling.

Significant overall estimates of Faith’s PD index revealed a decrease for BC cases (MD = -5.25; 95% CI = -6.35 to -4.15; p < 0.001) (Figure 7). There was no heterogeneity among studies (*I*^*2*^ = 0%; p = 0.52).

**Figure 7.**
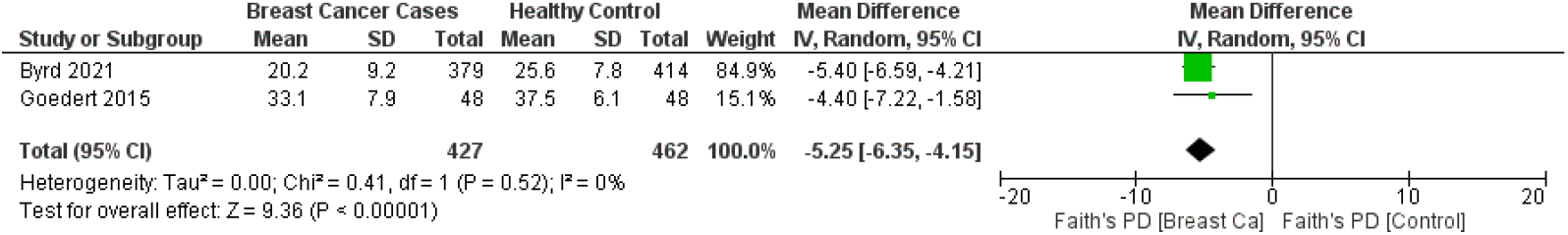
Meta-analysis forest plot representing the risk of breast cancer by faith’s PD of microbial profiling.

### Menopause

A study investigated the microbiome profile in fecal gDNA only on post-menopausal breast cancer women and control subjects, then the case and control study discovered the significantly altered microbial community of the cases (p = 0.006) compared with the controls and less alpha diversity (p = 0.004) (46).

### Menarche

It revealed that 46% of late menarche status has occurred in breast cancer cases (Table 2). In addition, the early and late menarche was associated with a low number of OTUs (p = 0.036), particularly reduced expression of Firmicutes (p = 0.048), and low chao1 index (p = 0.020) (72).

### Hormonal status

Goedert (2015) showed that a two-fold higher level of estrogen expression was found in post-menopausal patients, however, the difference did not change the microbiota and cancer association (46). Banerjee et al. (2018) found that an increased abundance of *Brevundimonas* was detected in ER^+^ BC and triple positive BC (TPBC) cases compared to the ER^-^ BC and TNBC. In addition, an abundance of *Mobiluncus* and *Mycobacterium* were predominantly identified in the ER^-^ BC samples. Furthermore, *Acinetobacter* was the most prominent in HR^+^ BC and HER2+ breast cancer cases, *Brevundimonas* in TPBC samples, and *Caulobacter* in TNBC samples (54). Interestingly, a distinct pattern of microbial profile in TNBC patients was explored using pan-pathogen array technology and described in the summary as in Table 3 (47).

**Table 3:**
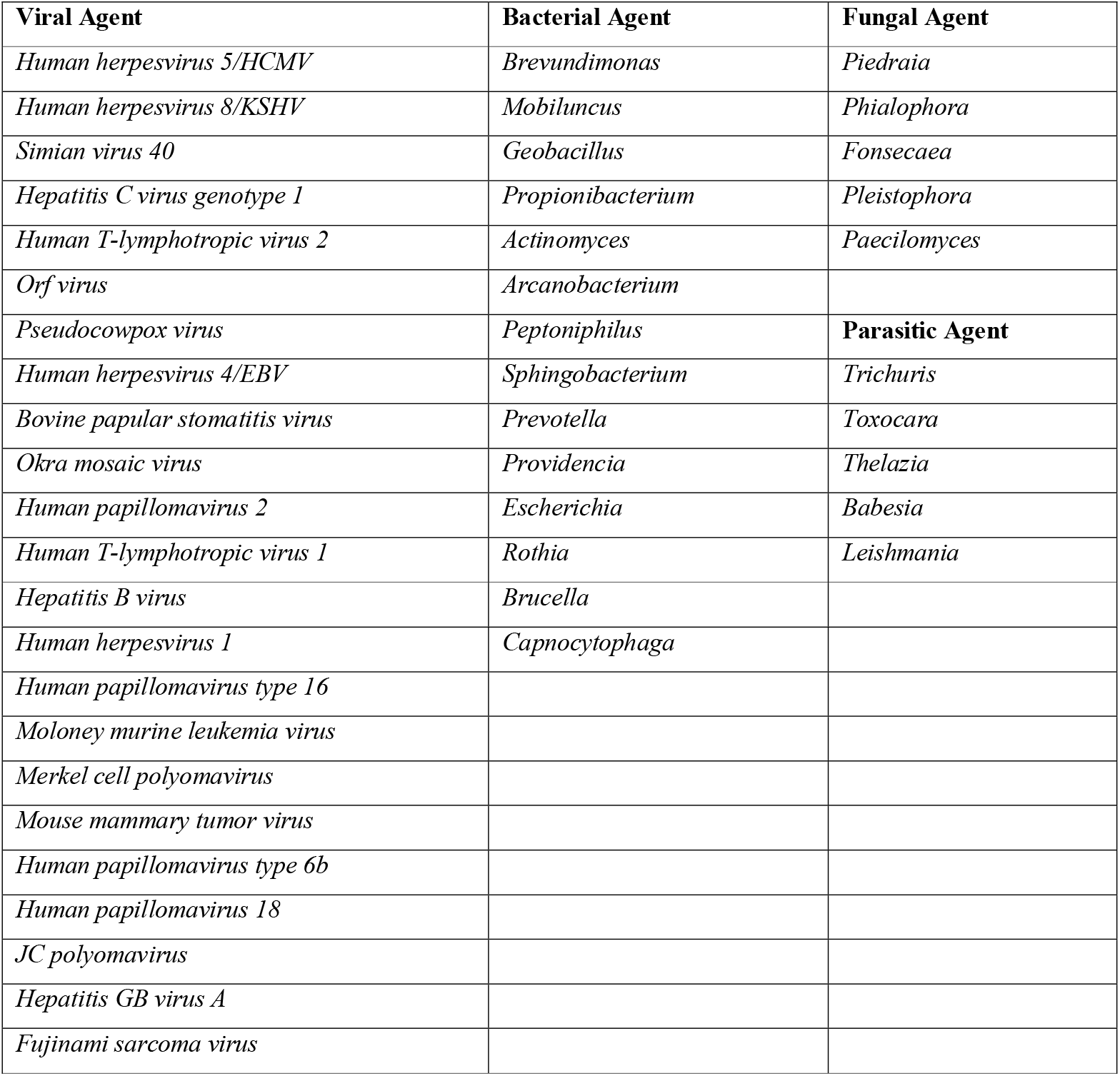
A distinct pattern of microbial profile in TNBC patients.

| **Viral Agent** | **Bacterial Agent** | **Fungal Agent** |
| --- | --- | --- |
| *Human herpesvirus 5/HCMV* | *Brevundimonas* | *Piedraia* |
| *Human herpesvirus 8/KSHV* | *Mobiluncus* | *Phialophora* |
| *Simian virus 40* | *Geobacillus* | *Fonsecaea* |
| *Hepatitis C virus genotype 1* | *Propionibacterium* | *Pleistophora* |
| *Human T-lymphotropic virus 2* | *Actinomyces* | *Paecilomyces* |
| *Orf virus* | *Arcanobacterium* |  |
| *Pseudocowpox virus* | *Peptoniphilus* | **Parasitic Agent** |
| *Human herpesvirus 4/EBV* | *Sphingobacterium* | *Trichuris* |
| *Bovine papular stomatitis virus* | *Prevotella* | *Toxocara* |
| *Okra mosaic virus* | *Providencia* | *Thelazia* |
| *Human papillomavirus 2* | *Escherichia* | *Babesia* |
| *Human T-lymphotropic virus 1* | *Rothia* | *Leishmania* |
| *Hepatitis B virus* | *Brucella* |  |
| *Human herpesvirus 1* | *Capnocytophaga* |  |
| *Human papillomavirus type 16* |  |  |
| *Moloney murine leukemia virus* |  |  |
| *Merkel cell polyomavirus* |  |  |
| *Mouse mammary tumor virus* |  |  |
| *Human papillomavirus type 6b* |  |  |
| *Human papillomavirus 18* |  |  |
| *JC polyomavirus* |  |  |
| *Hepatitis GB virus A* |  |  |
| *Fujinami sarcoma virus* |  |  |

## Discussion

The systematic literature review covered a long timeframe of 3 decades and nearly a dozen of countries’ data concerning the microbiome and BC. The meta-analysis revealed the microbial alteration in BC patients compared to control subjects and the microbial profile and diversity affected by a range of different BC treatments. Therefore, our study filled up the gap enabling us to link previous microbiome studies on BC patients through qualitative and quantitative meta-analysis tools.

The forest plots presented that the breast cancer patients have a lower alpha diversity compared with control subjects even though the Shannon index from a study is not significant.

Among multiple drivers of microbial differences, a common item is a menopausal status and we found that the BC patients with premenopausal status had an increased fecal profile of *Enterobacteriaceae*, aerobic *Streptococci, Lactobacilli* and anaerobic bacteria including *Clostridia, Bacteroides*, and *Lactobacilli* (42). Similar behavior of the anaerobic flora was found in late menopause patients. The urine microbiome from peri/post-menopausal patients also displayed a reduced abundance of *Lactobacilli* and elevated profile of many genera, comprising but not limited to anaerobic bacteria such as *Varibaculum, Porphyromonas, Prevotella, Bacteroides*, and members of the class Clostridia (50). More details of the microbiota in different groups were described in Tables 4-6.

**Table 4:** Characteristics of case-control studies and their microbial profiling and diversity.

| Ref. | Study Period | Sample | Age (Mean±SD) | Microbial detection method | Bacterial profile and diversity | Summary |
| --- | --- | --- | --- | --- | --- | --- |
| Minelli (1990) | - | Feces samples of 18 BC patients and 30 healthy subjects | - | Microbial cultures | Premenopausal patients: ↑ *Enterobacteriaceae* (*E. coli* and lactose non-fermenters, aerobic *streptococci*, and *lactobacilli*; anaerobes (*Clostridia*, *Bacteroides*, and *Lactobacilli*); No increase for anaerobic cocci.<br>Patients <5 years after Menopause: ↑ aerobic *Lactobacilli*, fungi, *E. coli*, *Bacteroides*, *Clostridia*.<br>Patients >5 years after Menopause: ↑ anaerobic *Lactobacilli*, *Bacteroides*, *Clostridia*. | The bacterial composition (both bacterial load and species) was different between BC and healthy women. |
| Benini (1992) | - | Feces samples of 28 BC patients and 45 healthy subjects | - | Microbial cultures | Premenopausal patients: ↑ *E. coli*, aerobic and anaerobic *Lactobacilli*, *Bacteroides*, and *Clostridia*<br>Patients <5 years after Menopause: ↑ *E. coli* and *Bacteroides*, ↓ *Enterococci*<br>Patients >5 years after Menopause: ↑ *E. coli*, *Bacteroides*, and *Clostridia*. ↓ Fungi. | The colonic flora of CA breast (both bacterial load and species) was significantly higher than that of healthy women.<br>A possible bacterial interference on sex steroid hormones metabolism and/or various accessibility of estrogens in BC were observed. |
| Xuan (2014) | - | 20 tumor tissue and 20 paired normal tissue | 63.43 | Pyrosequencing, Illumina | 96.6% of all samples are Proteobacteria, Firmicutes, Actinobacteria, Bacteroidetes, and Verrucomicrobia.<br>BC patients (100%, p = 0.015): ↑ *Methylobacterium radiotolerans* (66.67%, 2/3 OTUs)<br>Paired normal tissue (95%, p = 0.0097): | An inverse association of BC stage and bacterial load was observed in tumor tissue, but not in paired normal tissue.<br>A significant reduction in anti- |
|  |  |  |  |  | ↑ *Sphingomonas yanoikuyae* (50%, 4/8 OTUs) | bacterial responses was found in tumor tissue as a consequence of their impact on the local immune microenvironment in BC. |
| Urbaniak (2014) | 2012 | Breast tissue samples from Canadians including 27 BC, 11 benign, and 5 healthy subjects, and Irish accounting for 33 CA patients and 5 healthy. | - | Ion-Torrent sequencing | Canadian: *Bacillus* (11.4%), *Acinetobacter* (10.0%), *Enterobacteriaceae* (8.3%), *Pseudomonas* (6.5%), *Staphylococcus* (6.5%), *Propionibacterium* (5.8%), *Comamonadaceae* (5.7%), *Gammaproteobacteria* (5.0%), and *Prevotella* (5.0%)<br>Irish: *Enterobacteriaceae* (30.8%), *Staphylococcus* (12.7%), *Listeria welshimeri* (12.1%), *Propionibacterium* (10.1%), and *Pseudomonas* (5.3%) | A broad population of bacteria was found in tissue taken from several locations around the breast in women who did not all have a history of lactation.<br>The most common phylum was Proteobacteria. |
| Goedert (2015) | - | Feces samples of 48 BC patients and 48 controls | 62 | Illumina Sequencing | Except for the Shannon index ($p = 0.09$), the fecal microbiota of case patients had statistically substantially decreased α-diversity ($p \leq 0.004$) than that of control patients.<br>In healthy people, fecal microbiota diversity (PD whole tree) was found to be directly associated with total estrogens ($\rho = 0.37$, $p = 0.009$). They are unrelated to BC patients.<br>Across all genus-level taxa, there was a difference in fecal microbiota composition (β-diversity) between case and control patients (unweighted UniFrac $p = 0.009$). | Fecal microbiota in BC patients was less diverse and compositionally different, and they had increased levels of systemic estrogens. |
| Banerjee (2015) | - | FFPE tissue samples from 100 CA patients, 17 adjacent healthy tissue of the patients, and 20 healthy subjects. | - | Illumina Sequencing, Microarray | TNBC samples: Prevalent viral, bacterial, fungal, and parasitic genomic sequences were detected. | TNBC tissue had a distinct microbial profile that was not found in normal tissue.<br>Two broad microbiological fingerprints emerged from hierarchical clustering analysis, one common in bacteria and parasites and the other common in viruses. |
| Urbaniak (2016) | - | Fresh frozen tissue of 45 CA breasts, 13 benign patients, and 23 healthy controls. | 53.5 | Illumina Sequencing | CA breast: ↑ proportional abundances of *Bacillus*, *Enterobacteriaceae*, and *Staphylococcus* | Normal nearby tissue from Ca breast patients and tissue from controls had different bacterial profiles.<br>The microbial profiles of normal surrounding tissue and malignant tissue were identical. |
| Hieken (2016) | - | Fresh frozen tissue of 17 CA breast and 16 benign patients. | 60 | Illumina Sequencing | CA breast: ↑ genera *Fusobacterium*, *Atopobium*, *Gluconacetobacter*, *Hydrogenophaga*, and *Lactobacillus* | The microbiota of breast tissue, breast skin swabs, and buccal swabs were all found to be different from the microbiota of breast skin tissue. In CA breast tissue vs. benign breast disease, different microbial populations were discovered. |
| Wang (2017) | 2014-2016 | Breast tissue samples from 57 CA breast patients and 21 healthy subjects. | 52.82 | Illumina Sequencing | BC case: ↓ Relative abundance of *Methylobacterium*, ↑ family *Alcaligenaceae*.<br>↓ *Lactobacillus* in urine largely explained by menopausal status, with peri/postmenopausal women<br>↑ GPB including *Corynebacterium* (p < 0.01), *Staphylococcus* (p = 0.02), *Actinomyces* (p < 0.01), and *Propionibacteriaceae* (p < 0.01) in cancer, independent of menopausal status.<br>The microbiome of urine from peri/post-menopausal | Patients with those without breast cancer have different microbiota in their local breast and urinary system.<br>There were no significant differences in overall diversity or microbiome composition between invasive carcinoma and paired normal tissue in patients with |
|  |  |  |  |  | group: ↓ abundance of genus *Lactobacillus*, and ↑ abundances of numerous other genera, including but not limited to anaerobes such as *Varibaculum*, *Porphyromonas*, *Prevotella*, *Bacteroides*, and members of the class *Clostridia*. | invasive cancer.<br>↓ genus *Methylobacterium* in CA breast tissue and relatively ↑ GP bacteria in the urinary microbiome.<br>In terms of oral rinse, there are no major variations. |
| Thompson (2017) | - | Frozen tissue blocks of 668 CA breast and 72 healthy subjects. | - | Illumina Sequencing | Tumor tissue: Proteobacteria (48%), Actinobacteria (26.3%), Firmicutes (16.2%), Others (9.5%) from miscellaneous phyla; ↑ abundance for *S. pyogenes* and *L. rossiae*. *L. fleischmannii* and, to a lesser extent, *N. Subflava* were found. *H. influenza* was found to be associated with genes involved in tumor growth pathways.<br>Non-cancerous adjacent tissue: The abundance ranges from 0.5 to 19.3 %, though these species make up 85.64 % of the entire microbiota in breast tissue. | Proteobacteria were found in greater quantity in tumor tissues, while Actinobacteria were found in greater abundance in non-cancerous neighboring tissues. |
| Goedert (2018) | - | Feces samples of 48 BC patients and 48 controls | 62 | Illumina Sequencing | Sequencing revealed 1561 microbial taxa. In the IgA-positive microbiota, the reduced richness and diversity were more noticeable. | Cases showed significantly reduced α-diversity and altered composition of both their IgA-positive and IgA-negative intestinal microbiota after adjusting for estrogens and other factors. Compared to IgA-positive and IgA-negative gut microbiota, BC patients demonstrated substantial estrogen-independent correlations. IgA-negative bacteria had a slightly higher overall richness than IgA-positive bacteria. |
| Huang (2018) | 2006-2015 | cfDNA of 3 BC patients and 2 healthy subjects | - | Illumina Sequencing | *Acinetobacter johnsonii* *XBB1* and low levels of *Mycobacterium spp.* were discovered in all healthy females but were also found in an early BC (EOBC) patient, but large titers of bacterial cfDNA in EOBC patients were obtained from *Pseudomonas* or *Sphingomonas spp.* | Bacterial species were more diverse and more likely to be present at high levels in EOBC patients. The majority of microbial species found in healthy human bodies have modest amounts of cfDNA. *Acinetobacter* (notably *A. johnsonii* *XBB1*) cfDNA was found in healthy people, however, it could also be found in EOBC patients. |
|  |  |  |  |  |  | In the plasma of BC patients with severe symptoms, *Pseudomonas* and *Sphingomonas* cfDNA can be identified. |
| Banerjee (2018) | - | FFPE tissue samples from 148 BC patients, and 20 healthy subjects. | - | Illumina Sequencing, Microarray | Fungi: only 7 fungal families (*Aspergillus*, *Candida*, *Coccidioides*, *Cunninghamella*, *Geotrichum*, *Pleistophora*, and *Rhodotorula*).<br>Parasites: *Ancylostoma*, *Angiostrongylus*, *Echinococcus*, *Sarcocystis*, *Trichomonas*, and *Trichostrongylus* were found uniquely associated with TPBC.<br>*Balamuthia* signatures were associated significantly with BRHR samples, and that of *Centrocestus*, *Contracaecum*, *Leishmania*, *Necator*, *Onchocerca*, *Toxocara*, *Trichinella*, and *Trichuris* were detected significantly only with TNBC samples. | The TN and TP samples had unique patterns, however, the ER<sup>+</sup> and HER2<sup>+</sup> samples had comparable microbial signatures, according to hierarchical cluster analysis of their discovered microbial signatures. |
| Zhu (2018) | - | Feces samples of 62 breast cancer patients and 71 controls | 37.06 vs 35.52 (Pre)<br>57.45 vs 56.89 (Post) | Illumina Sequencing | There were significant differences in the relative abundance of 45 species between postmenopausal patients and postmenopausal controls; 38 species were enriched in postmenopausal patients, including *Escherichia coli*, *Klebsiella* sp_1_1_55, *Prevotella amnii*, *Enterococcus gallinarum*, *Actinomyces* sp. HPA0247, *Shewanella putrefaciens*, and *Erwinia* | In Ca breast patients, microbial diversity was higher than in controls.<br>Between premenopausal patients and controls, relative species abundance in the gut microbiota did not differ significantly. |
|  |  |  |  |  | *amylovora*, and 7 species were less abundant in postmenopausal patients, including *Eubacterium eligens* and *Lactobacillus vaginalis*. | The gut microbial community composition and functions changed between postmenopausal BC patients and healthy controls. |
| Smith (2019) | - | Fresh frozen tissue from 64 CA patients, 11 adjacent healthy tissue of the patients, and 8 healthy subjects. | 45 | Illumina Sequencing | Normal vs Tumor: ↓f_*Pseudomonadaceae* (p_Proteobacteria), f_*Sphingomonadaceae* (p_Bacteroidetes), and f_*Ruminococcaceae* (p_Firmicutes); ↑f_*Actinomycetaceae* (p_Actinobacteria)<br>Tumor: ↑*Clostridia*, *Bacteroidia*, *WPS_2*, f_*Ruminococcaceae* (LDA > 4)<br>Normal pairs: ↑f_*Pseudomonadaceae*, *Sphingomonadaceae*, and *Caulobacteraceae* (LDA > 5).<br>Stage 1: ↑p_Proteobacteria; f_*Ruminococcaceae* (p_Firmicutes), and g_*Hyphomicrobium* (p_Proteobacteria).<br>Stage 2: ↑p_Euryarchaeota, Firmicutes, and Spirochaetes; g_*Sporosarcina* (p_Firmicutes)<br>Stage 3/4: ↑p_Thermi, Gemmatimonadetes, and Tenericutes; g_*Bosea* (p_Proteobacteria) | Proteobacteria was most abundant but Firmicutes, Bacteroidetes, and Actinobacteria were the least numerous in all subjects.<br>When compared to non-Hispanic white tumors, non-Hispanic black breast tissues had a greater abundance of g_*Ralstonia*.<br>With each stage, the abundance of g_*Bosea* rose. |
|  |  |  |  |  | Luminal B tumors: ↑ g_Clostridium (p_Firmicutes);<br>Luminal A tumors: ↑ o_Xanthomonadales (p_Proteobacteria) (LDA > 5);<br>HER2 tumors: ↑ g_Akkermasia (p_Verrucomicrobia) (LDA = 4)<br>TNBC: f_Streptococcaceae in TNBC; also g_Streptococcaceae, g_Ruminococcus (both p_Firmicutes) (LDA > 3.5) |  |
| Ma (2020) | 2017-2018 | Feces samples of 25 breast cancer patients, and 25 benign patients as controls | - | Illumina Sequencing | CA breast: ↓ Relative abundance of *Firmicutes* and *Bacteroidetes*; ↑ relative abundance of Verrucomicrobiota, Proteobacteria, and Actinobacteria; ↓ Abundance of *Faecalibacterium* | The IL-6/STAT3 pathway was inhibited by *Faecalibacterium prausnitzii*, which inhibited the development of BC cells. |
| Klann (2020) | - | Breast tissue samples of 10 BC patients and 36 healthy samples. | - | Illumina Sequencing | Most abundant phyla are Bacteroidetes, Firmicutes, Proteobacteria, and Actinobacteria.<br>Most abundant OTUs: f_*Ruminococcaceae* and g_*Acidaminococcus*, *Acinetobacter*, *Akkermansia*, *Bacteroides*, and *Sutterella*. | There were significant differences in tumor and normal tissue variety, as well as in composition between women and between breasts of the same woman. |
| Uzan-Yulzari (2020) | - | Feces samples of 28 BC and 5 benign cancer patients | - | Illumina Sequencing | In the pretreatment microbiomes of women who gained weight following chemotherapy, LEfSe analysis revealed a higher relative abundance of members of the f_ *Erysipelotrichaceae* (and at the class and order levels as well).<br>β-diversity revealed a significant differences between the 2 groups.<br>Significant differences in α-diversity were found, indicating that the weight gain group had a more diversified pretreatment microbiome. | Weight gain after adjuvant chemotherapy in women with breast and gynecological malignancies was connected to the composition and diversity of the gut microbiome. |
| He (2021) | 2019 | Feces samples of 54 BC patients and 28 controls | 39.74 vs 37.54 | Illumina Sequencing | Firmicutes/Bacteroidetes (F/B) ratio was substantially higher in BC than that in controls.<br>The most abundant flora are Firmicutes, Proteobacteria, Bacteroidetes, and Actinobacteria<br>CA Breast: ↓ p_Acidobacteria, Nitrospirae, Fusobacteria, and Cyanobacteria/Chloroplast; ↑ *Synergistetes*; At the species level, the top 10 bacteria that decreased significantly are *Allisonella*, *Megasphaera*, *Pediococcus*, *Abiotrophia*, *Granulicatella*, and *Clostridium_sensu_stricto* belonging to Firmicutes, *Serratia* and *Enhydrobacter* belonging to Proteobacteria, *Fusobacterium* belonging to Fusobacteria, and *Slackia* belonging to | Premenopausal breast cancer patients and normal premenopausal women could be distinguished by *Pediococcus* and *Desulfovibrio*.<br>Pre-menopausal BC vs Normal:<br>The composition and symbiosis of intestinal flora changed significantly. |
|  |  |  |  |  | Actinobacteria. |  |
| Byrd (2021) | - | Feces samples of 379 CA breast, 102 benign patients (NM), and 414 healthy controls (NC). | CA: 50.8<br>NM: 38.8<br>NC: 46.9 | Illumina Sequencing | α-diversity metrics are strongly, inversely associated with the odds of breast cancer and for those in the highest vs. lowest tertile of observed ASVs, the odds ratio (95% CI) was 0.21 (0.13–0.36; $p < 0.001$).<br>It is similar for nonmalignant breast disease and breast cancer grade/ molecular subtype.<br>β-diversity distance matrices and multiple taxa with possible estrogen-conjugating and immune-related functions are strongly associated with CA breast. | Characteristics of feces bacteria were strongly and similarly linked to BC and non-malignancy. |

**Table 5:**
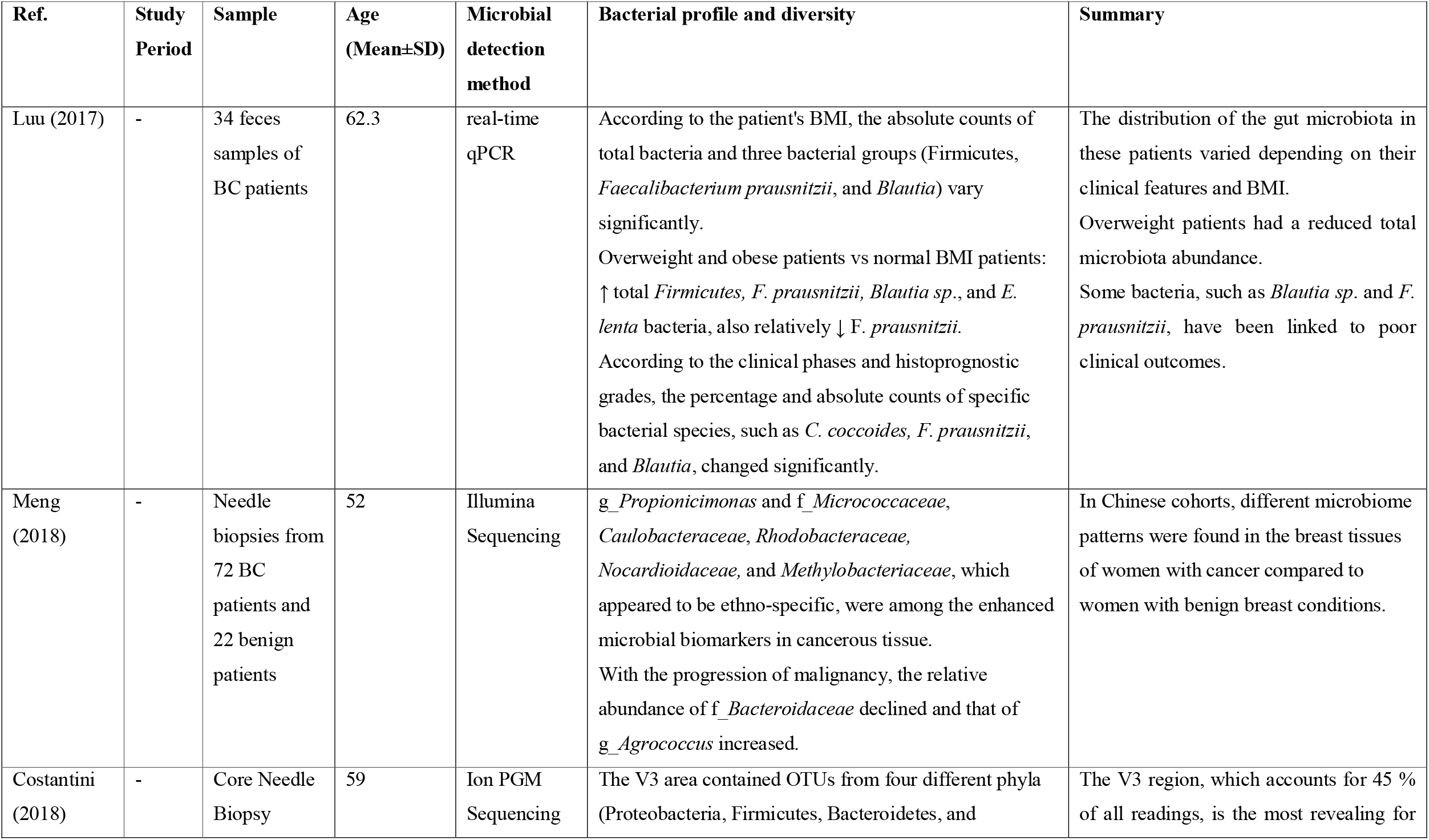
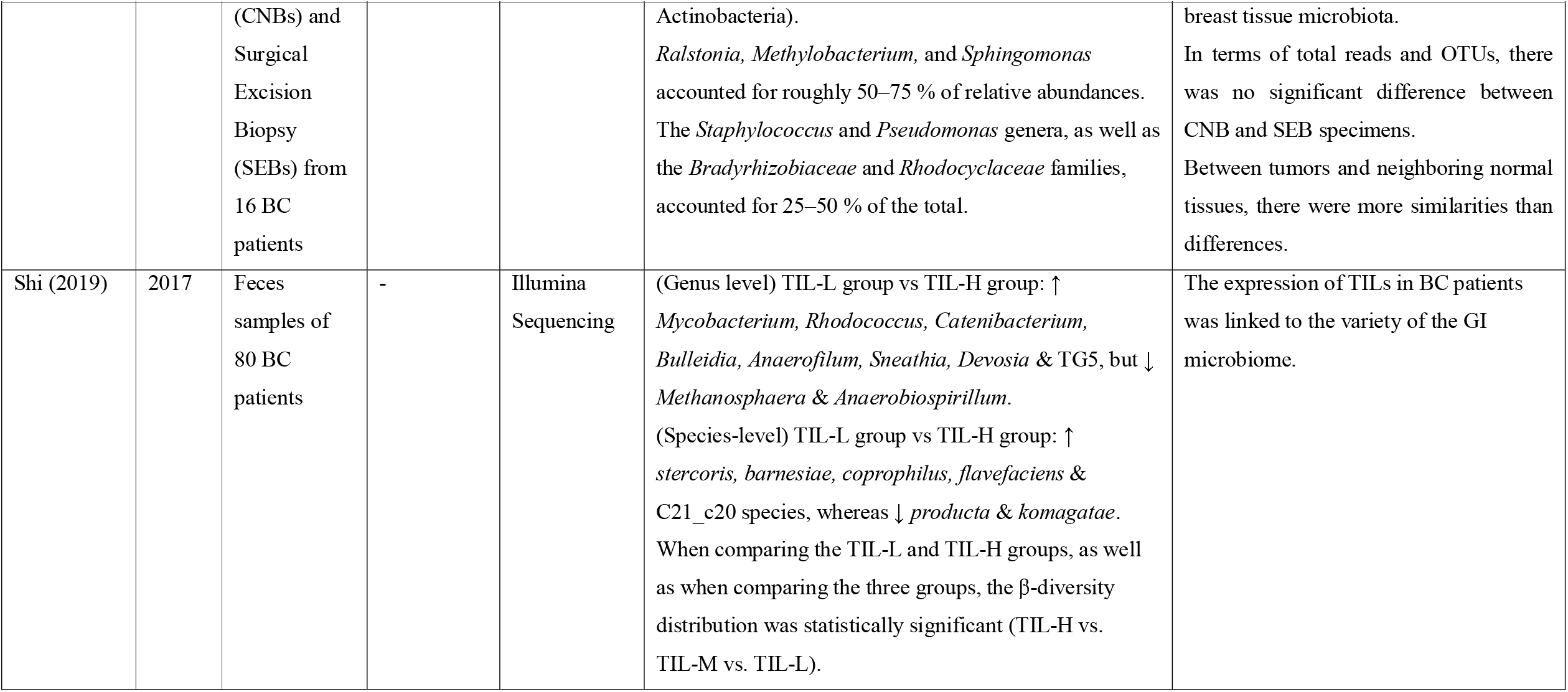
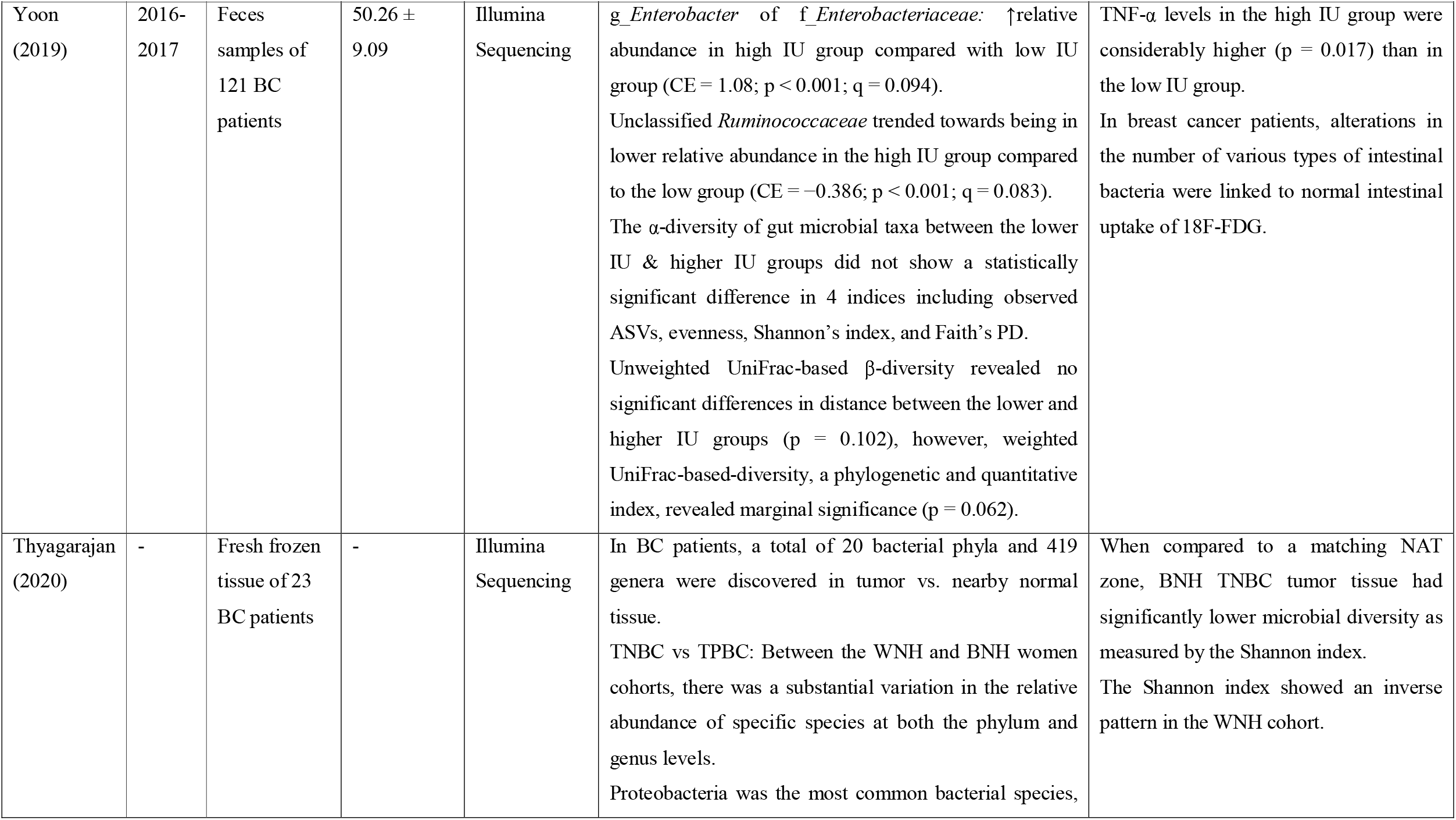
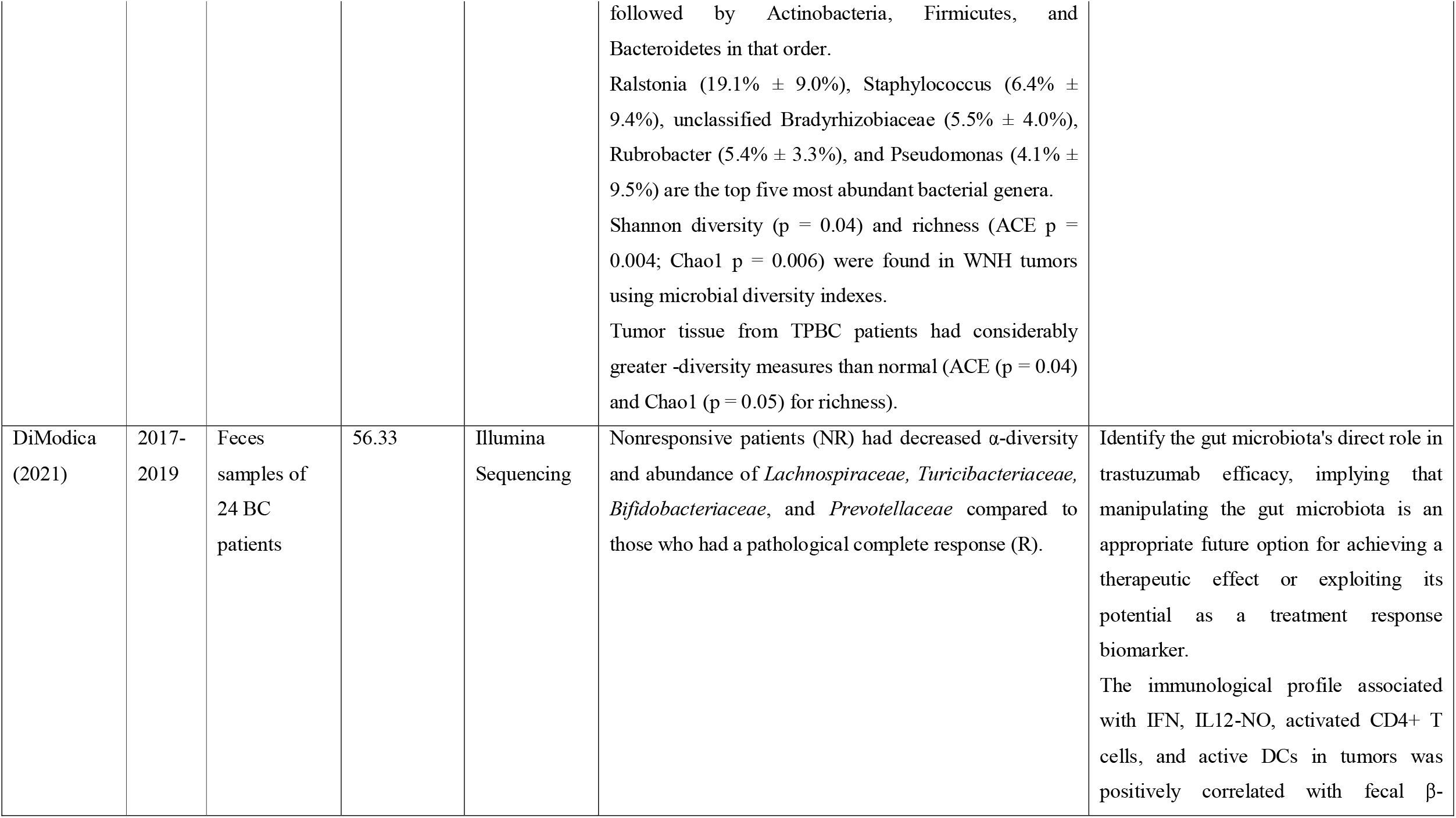
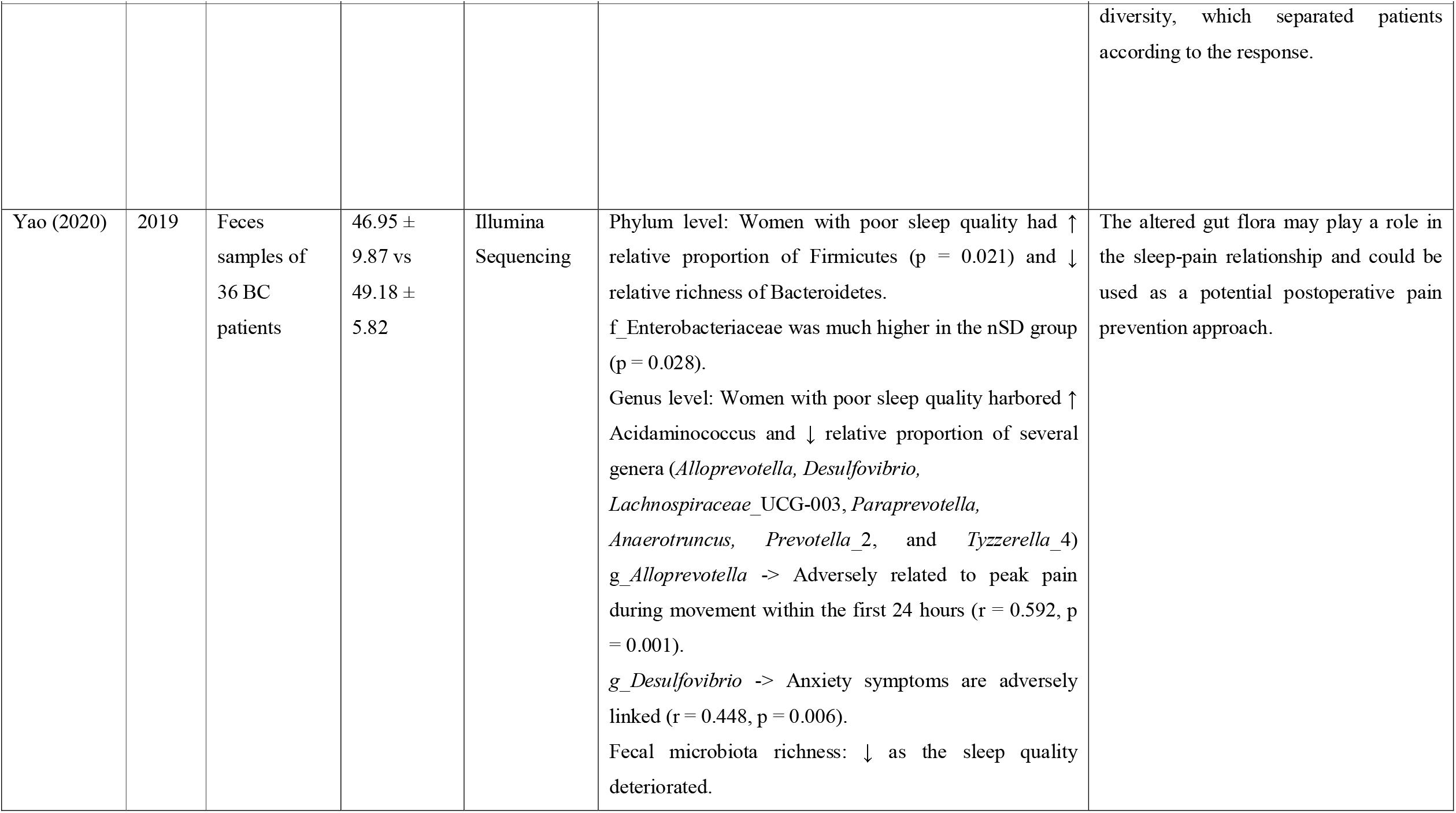
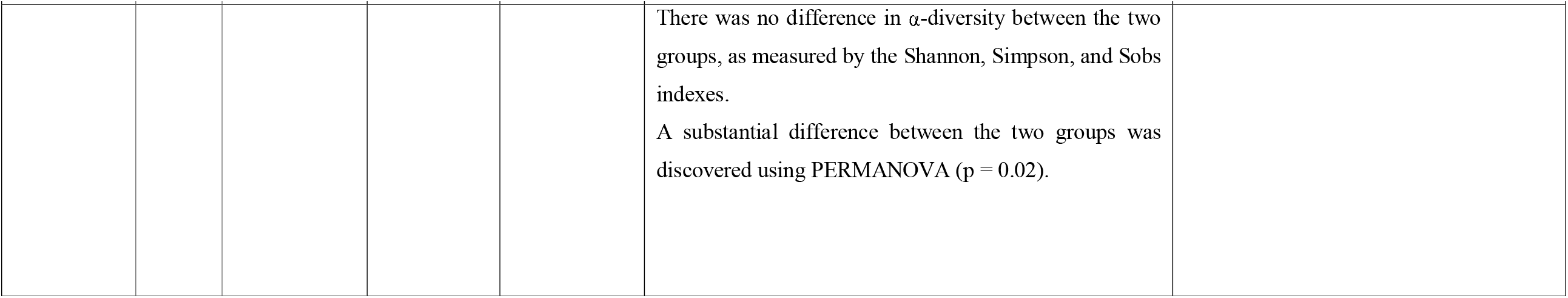
Characteristics of cohort studies and their microbial profiling and diversity.

**Table 6:**
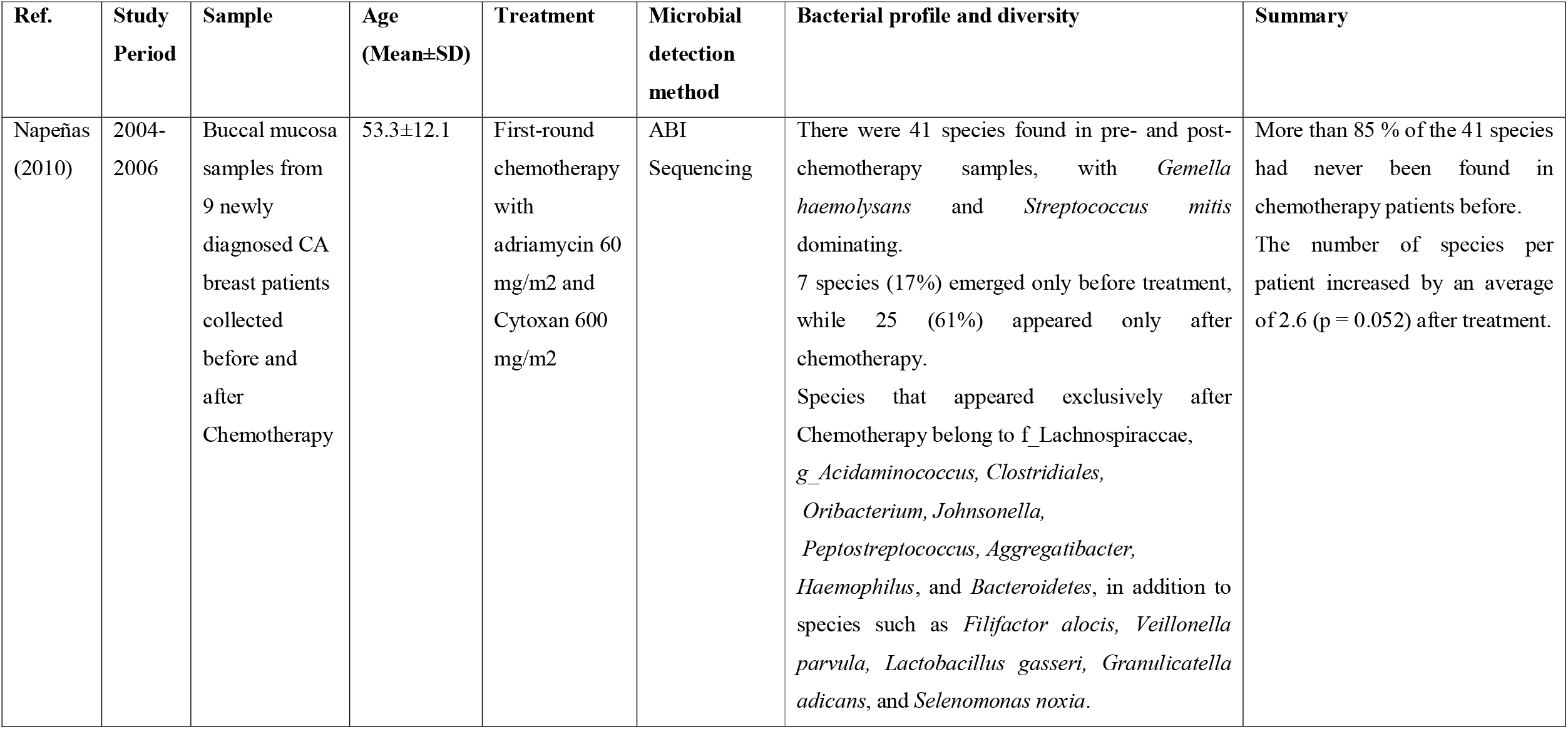
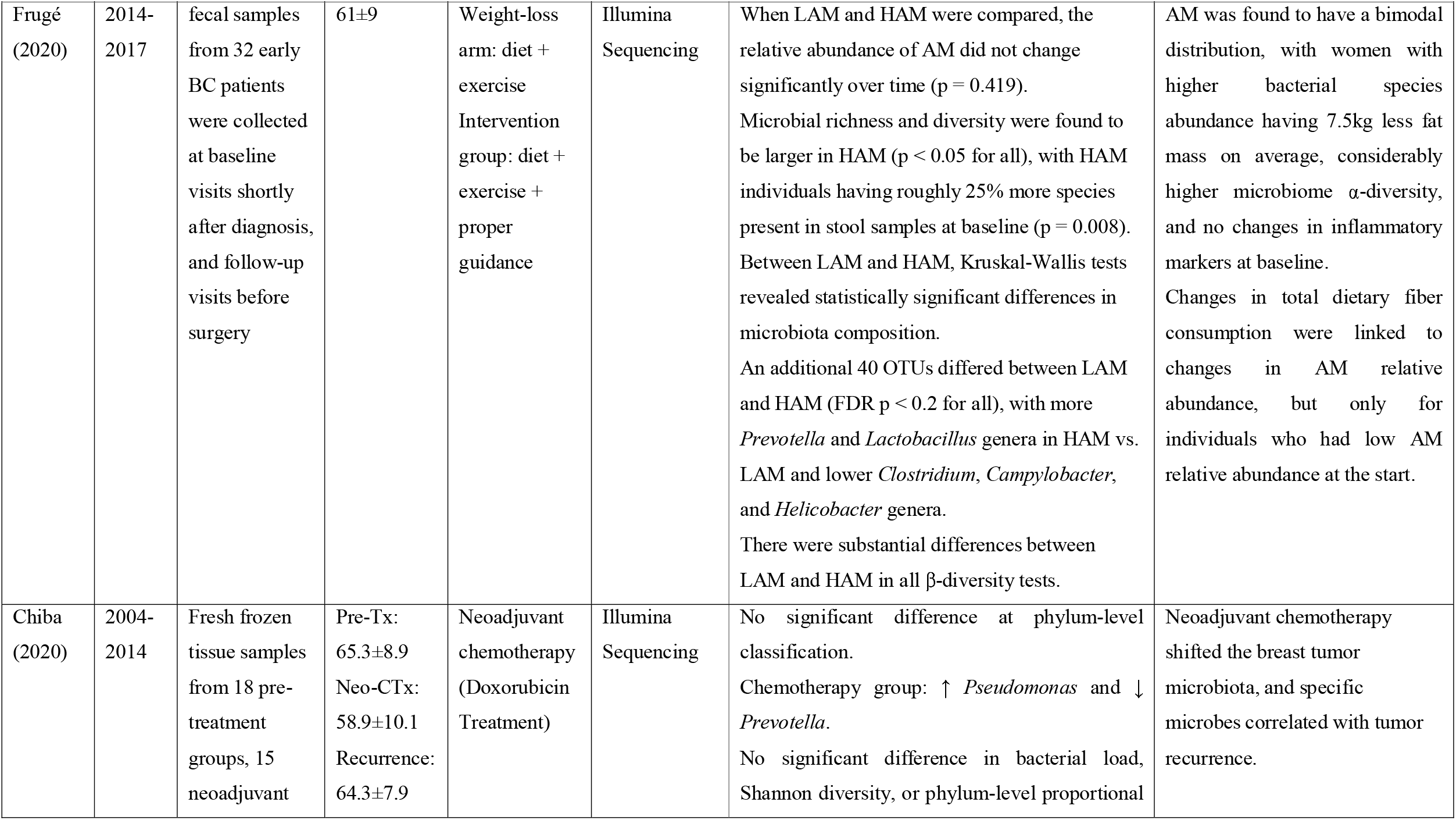
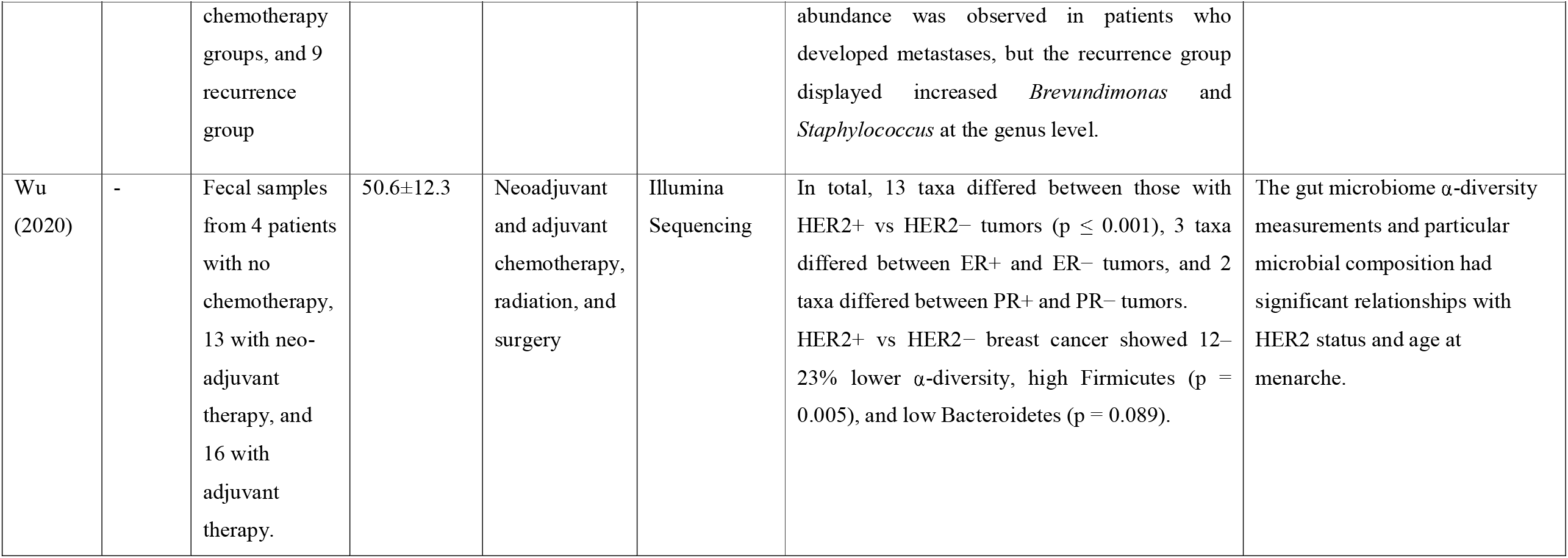
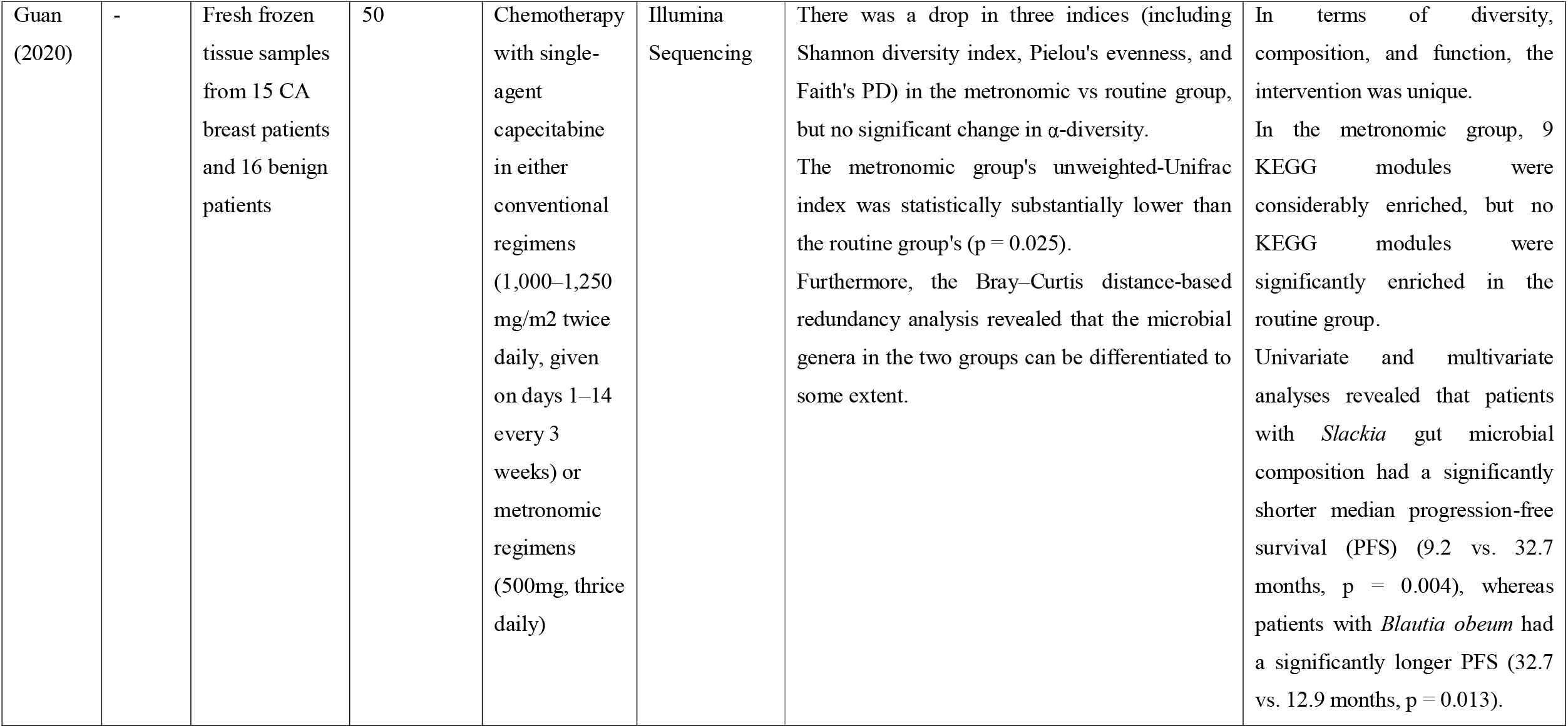
Characteristics of non-randomized intervention trials and their microbial profiling and diversity.

Even though the role of the species in the equilibrium of the GI environment is not clear, the increased bacterial concentration can modulate estrogen metabolism via deconjugation and contribute to total bacterial enzyme activity (42). As an example, the enzymes β-glucoronidase and β-glucosidase are produced by *E. coli* and *S. fecalis*, respectively. From an observation study of the enzymatic activity of fecal bacteria, it was acknowledged that the activity in post-menopausal women was lower than that of pre-menopausal cases. The expression of enzymes from high to low such as esterase-C4, esterase-lipase, leucine- and valine acrylamidase, acid phosphatase, α-galactosidase, and β-glucoronidase, were found in healthy subjects while leucine- and valine acrylamidase, β-glucoronidase, and esterase-lipase were higher in post-menopausal BC women (43). Therefore, it showed an inter-subject variability for enzymatic activities.

From the quantitative analysis of microbiota, Xuan et. al, found a 10-fold elevation of bacteria in breast tumors compared with paired normal tissue, however, there was no significant difference between the latter and healthy breast tissue. Moreover, it revealed an inverse correlation between bacterial count in the tumor tissue and the breast cancer stage, showing the lowest 16S ribosomal DNA (rDNA) copy numbers in stage 3 patients (44). In addition, a distinct breast tissue microbiota was found in different breast skin tissue, breast skin swabs, and buccal swabs, and profound microbial communities between benign and malignant breast disease as well (49). Furthermore, it seems to have a geographical difference between the Canadian and Irish breast tissue microbiome, but it still needs more pieces of evidence to prove that (49). Also, the study proved a distinct profile of breast tissue microbiome with increased species richness compared to overlying skin tissue suggesting that the differences may be due to the difference in their environment and ecosystem (49).

A study also explored the expression level of antibacterial response genes in tumor tissue, paired normal tissue and health tissue, and it found that one-third of antibacterial genes were significantly downregulated in breast tumor cases after normalizing with a housekeeping gene β-actin, interestingly no more upregulated genes there (44). Furthermore, a significant reduction of the transcripts of microbial sensors TLR2, TLR5, and TLR9 (p = 0.0298, p = 0.0201, and p = 0.0021, respectively) was encountered in the tumor tissue while there was a similar expression level of TLR1, TLR4 and TLR6 in healthy and tumor tissue. In addition, tumor tissues displayed significantly decreased expression of cytoplasmic microbial sensors (NOD1 and NOD2), as well as downstream signaling molecules for innate microbial sensors such as CARD6, CARD9, and TRAF6 (p = 0.0207, p = 0.0040, and p = 0.0119, respectively). The levels of BPI, MPO, and PRTN3 are significantly decreased (p = 0.0133, 0.002, and 0.0022, respectively) (43). Even though further research is needed to confirm the influence of the breast tissue’s local microenvironment, these findings demonstrated a significant decrease in antimicrobial responses in breast tumor tissue. (44).

To explore the impact of cancer chemotherapy, Napenas et. al., performed a profiling of the oral microbiome on 9 newly diagnosed breast cancer patients before and after receiving the treatment (69). Overall, it detected 41 species in total, and interestingly, >85% of the detection (33/41) were the newly identified species in chemotherapy patients. It revealed that 7 species and 25 species appeared solely before and after the cancer chemotherapy respectively, and species increment per patient had a mean of 2.6 (SD = 4.7, p = 0.052) after chemotherapy (69).

Chiba et. al., evaluated the modulation of tumor microbiome by neoadjuvant chemotherapy using breast tumor microarrays (71). It demonstrated that there were no significant changes in total bacterial load in untreated and treated patients, and the bacterial diversity was significantly reduced in the treated tumor. The phylum-level classification displayed no significant changes between the two groups, but the genus-level analysis showed a significant elevation of *Pseudomonas* species and a reduction of *Prevotella* abundance in the treated cases (71). In addition, it indicated the modulation of chemotherapy on the tumor microbiome and the correlation of some genera in patients with tumor recurrence (71).

However, the meta-analysis study has some limitations. Firstly, several matrices for the detection of alpha diversity were utilized in different studies, and probably no standardized tools for the measurement. Therefore, only a few studies were able to analyze quantitatively. Secondly, some mean values cannot be found in the papers and supplementary files, and the email contacts for 2 weeks are reachable only to some, perhaps the contacts are changed, or the data are not archived for the long period. Hence, qualitative analysis was applied to the provided data of the articles.

Overall, our meta-analysis suggests the fecal, tumor, or oral microbiome profile of breast cancer patients, the differences in the abundance of microbiota by menopausal status, menarche, and cancer stages, and the microbial pattern changing before and after the chemotherapy. However, the investigation of the microbiome is still in the infancy for breast cancer patients and normally the sample size is limited due to high sequencing cost. Therefore, further studies with a larger cohort of patients would be required to identify the biological and pathological significance of the findings in the meta-analysis. We expected the review could fill up the gap linking to understand more connection between breast cancer and microbiome.

## Supporting information

PRISMA Checklist

Graphical Abstract

## Data Availability

All data produced in the present work are contained in the manuscript.

