## Supplementary A for "Microbiome and breast cancer: A systematic review and meta-analysis"

**Supplementary File 1. Full search strategy**

| **Set #** | **PubMed** | **Results** |
| --- | --- | --- |
| 1  Microbiome | "microbiota"[MeSH Terms] OR "gastrointestinal microbiome"[MeSH Terms] OR "mycobiome"[MeSH Terms] OR microbiome*[tiab] OR microbiota*[tiab] OR microbial[tiab] OR microbe*[tiab] OR microflora*[tiab] OR flora*[tiab] OR microorganism*[tiab] OR pathobiont*[tiab] OR mycobiome*[tiab] OR mycobiota*[tiab] OR virome*[tiab] OR phylotype*[tiab] OR enterotype*[tiab] | **403932** |
| 2  Breast Cancer | "breast neoplasms"[MeSH Terms] OR (breast[tiab] AND (cancer*[tiab] OR neoplas*[tiab] OR tumor*[tiab] OR tumour*[tiab] OR malignan*[tiab] OR carcinoma*[tiab] OR adenocarcinoma*[tiab])) | **429035** |
| 3 | #1 AND #2 | **945** |
| 4 | animals[MeSH Terms] NOT humans[MeSH Terms] | **4805329** |
| 5 | #3 NOT #4 | **896** |

| **Set #** | **Embase** | **Results** |
| --- | --- | --- |
| 1  Microbiome | 'microbiome'/exp OR 'microflora'/exp OR 'intestine flora'/exp OR 'mycobiome'/exp OR 'microbiome*':ti,ab OR 'microbiota*':ti,ab OR 'microbial':ti,ab OR 'microbe*':ti,ab OR 'microflora*':ti,ab OR 'flora*':ti,ab OR 'microorganism*':ti,ab OR 'pathobiont*':ti,ab OR 'mycobiome*':ti,ab OR 'mycobiota*':ti,ab OR 'virome*':ti,ab OR 'phylotype*':ti,ab OR 'enterotype*':ti,ab | **490885** |
| 2  Breast Cancer | 'breast cancer'/exp OR ('breast':ti,ab AND ('cancer*':ti,ab OR 'neoplas*':ti,ab OR 'tumor*':ti,ab OR 'tumour*':ti,ab OR 'malignan*':ti,ab OR 'carcinoma*':ti,ab OR 'adenocarcinoma*':ti,ab)) | **642120** |
| 3 | #1 AND #2 | **1947** |
| 4 | [animals]/lim NOT [humans]/lim | **5945936** |
| 5 | #3 NOT #4 | **1775** |

| **Set #** | **CENTRAL** | **Results** |
| --- | --- | --- |
| 1  Microbiome | [mh microbiota] OR [mh "gastrointestinal microbiome"] OR [mh mycobiome] OR microbiome*:ti,ab,kw OR microbiota*:ti,ab,kw OR microbial:ti,ab,kw OR microbe*:ti,ab,kw OR microflora*:ti,ab,kw OR flora*:ti,ab,kw OR microorganism*:ti,ab,kw OR pathobiont*:ti,ab,kw OR mycobiome*:ti,ab,kw OR mycobiota*:ti,ab,kw OR virome*:ti,ab,kw OR phylotype*:ti,ab,kw OR enterotype*:ti,ab,kw | **17075** |
| 2  Breast Cancer | [mh "breast neoplasms"] OR (breast:ti,ab,kw AND (cancer*:ti,ab,kw OR neoplas*:ti,ab,kw OR tumor*:ti,ab,kw OR tumour*:ti,ab,kw OR malignan*:ti,ab,kw OR carcinoma*:ti,ab,kw OR adenocarcinoma*:ti,ab,kw)) | **38735** |
| 3 | #1 AND #2 | **61** |
