## Supplementary figures and images for "Microbiome and breast cancer: A systematic review and meta-analysis"

### Graphical Abstract

**Graphical Abstract**

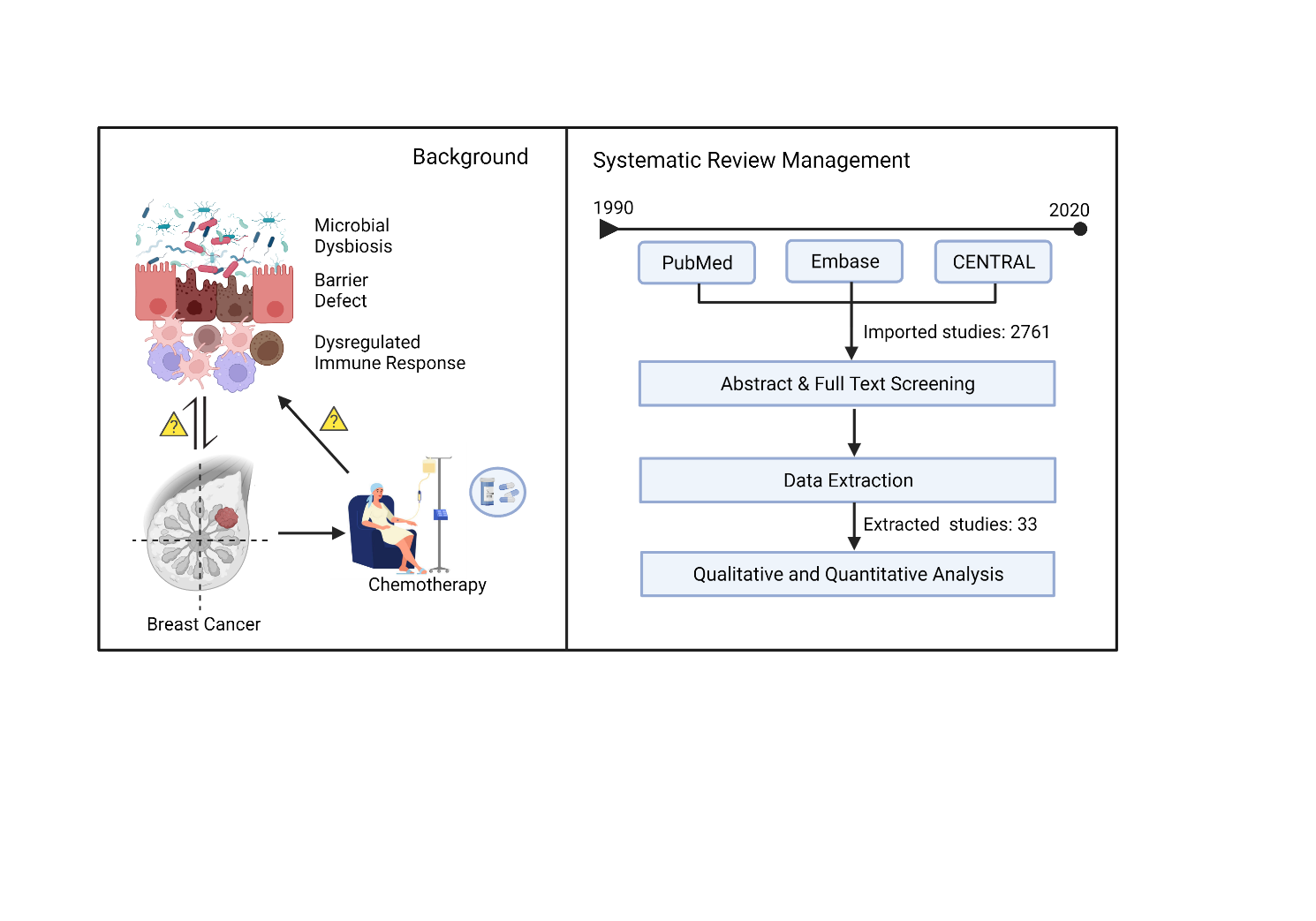
